# Multicentre accuracy trial of FUJIFILM SILVAMP TB LAM test in people with HIV reveals lot variability

**DOI:** 10.1101/2022.09.07.22278961

**Authors:** Rita Székely, Bianca Sossen, Madalo Mukoka, Monde Muyoyeta, Elizabeth Nakabugo, Jerry Hella, Hung Van Nguyen, Sasiwimol Ubolyam, Kinuyo Chikamatsu, Aurélien Macé, Marcia Vermeulen, Chad M Centner, Sarah Nyangu, Nsala Sanjase, Mohamed Sasamalo, Huong Thi Dinh, The Anh Ngo, Weerawat Manosuthi, Supunnee Jirajariyavej, Satoshi Mitarai, Nhung Viet Nguyen, Anchalee Avihingsanon, Klaus Reither, Lydia Nakiyingi, Andrew D. Kerkhoff, Peter MacPherson, Graeme Meintjes, Claudia M Denkinger, Morten Ruhwald, FujiLAM Study Consortium

## Abstract

**Rationale:** There is an urgent need for rapid, non-sputum point-of-care diagnostics to detect tuberculosis.

**Objectives:** This prospective trial in seven high tuberculosis burden countries set out to evaluate the diagnostic accuracy of the point-of-care urine-based lipoarabinomannan assay FUJIFILM SILVAMP TB LAM (FujiLAM) among inpatient and outpatient people living with HIV.

**Methods:** Diagnostic performance of FujiLAM at point of care was assessed among adult people with HIV against a mycobacterial reference standard (sputum culture, blood culture, and Xpert Ultra from urine and sputum at enrollment, and additional sputum culture ≤7 days from enrollment), an extended mycobacterial reference standard including available non-study test results, and a composite reference standard including clinical evaluation.

**Measurements and Main Results:** Of 1624 participants enrolled, 294 (18·0%) were classified as TB positive by eMRS. Median age was 40 years, median CD4 cell count was 372 cells/ul, 52% were female and 78% were taking antiretroviral therapy at enrollment. Overall FujiLAM sensitivity was 54·8% (95% CI: 49·1–60·4), and overall specificity was 85·1% (83·1–86·9), against the extended mycobacterial reference standard. Sensitivity and specificity estimates varied between sites, ranging from 26·5% (95% CI: 17·4%–38·0%) to 83·3% (43·6%–97·0%), and 75·0 (65·0%–82·9%) to 96·5 (92·1%–98·5%), respectively. Post-hoc exploratory analysis identified significant variability in the performance of the six FujiLAM lots used in this study.

**Conclusions:** Lot variability limited interpretation of FujiLAM test performance. Although the results with the current version of FujiLAM are too variable for clinical decision-making, the lipoarabinomannan biomarker still holds promise for tuberculosis diagnostics.

## Introduction

Tuberculosis (TB) is a leading cause of death from a single infectious disease, second only recently to COVID-19 (1). In 2020, TB caused 1·5 million deaths, including 214,000 among people living with HIV (PLHIV) (2). TB is the most common cause of death in PLHIV, who have a 30-times greater risk of developing TB disease than those without HIV (3). Most of the deaths from TB would be preventable if TB were diagnosed earlier, yet TB often goes undiagnosed (4-6).

Traditional diagnostic methods for TB, such as culture or smear microscopy, are slow or have low sensitivity. More sensitive modern techniques, such as Xpert^®^ MTB/RIF (Cepheid, Sunnyvale, CA, USA), require some laboratory infrastructure, are costly, and often inaccessible at the primary healthcare level where people at greatest risk of TB disease often first seek care. Moreover, TB is harder to diagnose in PLHIV, who frequently have paucibacillary, extrapulmonary or disseminated TB, and often experience difficulty producing sputum specimens (7, 8). TB in PLHIV is associated with high mortality if undiagnosed or if treatment is delayed (9). New, rapid, non-sputum-based point-of-care (POC) diagnostic tests to detect TB are urgently needed (10).

FUJIFILM SILVAMP TB LAM (FujiLAM; Fujifilm, Tokyo, Japan) is a visually read, qualitative, rapid, in vitro diagnostic test for the detection of the lipoarabinomannan (LAM) antigen of *Mycobacterium tuberculosis* (MTB) in human urine (11). FujiLAM includes two monoclonal antibodies, which bind to glycan capping motifs of LAM, and a silver amplification immunochromatography step, which enables an approximately 30-fold lower limit of detection compared with conventional lateral flow immunoassays (e.g. the Determine™ TB LAM Ag “AlereLAM”, Abbott, Chicago, IL, USA) (11, 12). The binding targets of glycan capping motifs also result in increased specificity for MTB complex (13).

In a study of frozen urine samples from inpatient PLHIV, FujiLAM showed superior diagnostic sensitivity (70% vs. 42%) with similar specificity (91% vs. 95%) to AlereLAM (11, 14). Here, we report results from the first large-scale, multicentre evaluation of FujiLAM accuracy on prospectively collected, fresh urine samples from PLHIV against a comprehensive reference standard.

## Methods

### Study design and participants

This was a prospective, multicentre cohort study, with consecutive patient recruitment from clinical sites in seven high TB burden countries (Malawi, South Africa, Tanzania, Thailand, Uganda, Viet Nam and Zambia). Participating centres are described in Table E1 in the online data supplement. The study recruited adult (≥18 years) PLHIV, irrespective of CD4 counts and antiretroviral therapy (ART) status, who had received no or <3 doses of anti-TB treatment in the last 60 days and no isoniazid preventive therapy within the 6 months prior to enrollment. Patients recruited from outpatient settings were included if they had at least one symptom suggestive of TB (current cough, night sweats, fever, weight loss); inpatients were enrolled irrespective of TB symptoms.

To assess the diagnostic accuracy of FujiLAM, multiple reference standards were used: a microbiological reference standard (MRS), extended MRS (eMRS), and composite reference standard (CRS) as per definitions in Supplementary Table E2.

The primary objectives of the study were to determine the diagnostic accuracy of FujiLAM for TB detection among PLHIV against the eMRS and CRS.

Participants were invited to provide samples on the day of enrollment (Day 1) and again within 7 days of enrollment (Day 2). All participants with negative eMRS results at baseline were followed up 2–3 months after enrollment, and an additional sample was collected if signs/symptoms (e.g. cough, fever) had not improved or completely resolved compared with baseline, as assessed by the local provider. Patients with baseline FujiLAM-positive but negative CRS results were invited to come back at 6 months, when additional samples were collected if signs/symptoms had not improved or completely resolved.

All study-related activities were approved by each country’s Research Ethics Committee. Written informed consent was obtained from participants, as per the study protocol. Study participation did not affect standard of care. The full study protocol is available at clinicaltrials.gov (NCT04089423).

### Procedures

The testing flow and number of samples tested are shown in Figure E1 in the online data supplement.

Urine, sputum and blood specimens were collected and processed fresh from participants after informed consent was obtained and clinical assessment was completed. Urine specimens were collected on the day of enrollment (spot urine), within 7 days of enrollment (early morning) and at the 6-month follow-up visit (if indicated). Urine samples were tested using FujiLAM and AlereLAM, and remaining urine samples were submitted for Xpert MTB/RIF Ultra (Cepheid, Sunnyvale, CA, USA) testing. For this, 30 ml urine was centrifuged at 3000x g for 15 minutes and, following removal of the supernatant, the pellet was re-suspended in 0.75 ml phosphate-buffered saline and 1·5 ml sample reagent buffer. Subsequently, 2 ml of the reagent-treated specimen was tested. When possible, left-over urine was preserved at _J80 _JC on-site for additional testing.

Blood was collected on Day 1 and submitted for CD4 cell count (flow cytometry) and mycobacterial blood culture. Sputum samples were collected on study Days 1 (spot) and 2 (early morning), and at the 3- and 6-month visits (if indicated) and tested by smear microscopy (fluorescence microscopy using Auramine O staining and/or Ziehl-Neelsen staining), Mycobacteria Growth Indicator Tube liquid culture (MGIT; Becton Dickinson, Franklin Lakes, NJ, USA), solid culture on Löwenstein-Jensen (LJ) medium, and Xpert MTB/RIF Ultra. If a participant was unable to provide sputum spontaneously, an attempt was made to collect induced sputum (depending on site regulations, participant health status, and COVID-19 restrictions).

Speciation was done from any positive mycobacterial culture (sputum, blood) using MPT64 antigen detection and/or MTBDR*plus*, MTBC and CM/AS line probe assays (Hain Lifescience, Nehren, Germany). Blood culture from all participants were done in BACTEC™ Myco/F Lytic culture vials (Becton Dickinson, Franklin Lakes, NJ, USA). WHO prequalified in vitro rapid diagnostic tests were used for HIV testing. Chest X-ray was performed on the day of enrollment if not done already by the treating clinical team. Day 1 samples were collected on the day of enrollment and tested within 24 hours; Day 2 samples were collected within seven days of enrollment and tested within 24 hours. Additional non-study samples were collected at the discretion of the treating clinician.

#### FujiLAM testing

Testing with the investigational product, FujiLAM, was performed at POC (either bedside/adjacent room a few meters from the ward/clinic) according to manufacturer’s instructions on the day of collection (11, 15); ideally within 2 hours of sample collection, or samples were kept at 2–8 °C until testing occurred.

#### AlereLAM testing

AlereLAM testing was performed at POC as per manufacturer’s instructions, using the test’s Reference Scale Card of 4 grades with the Grade 1 cut-off point as the positivity threshold.

AlereLAM and FujiLAM tests were done for each patient at Day 1, Day 2, and 6-months follow-up (if indicated). Both tests were performed and interpreted independently by different operators to ensure blinding. Operators were also blinded to reference tests and all results obtained from other tests. Laboratory personnel performing the reference standard testing did not have access to the results of the FujiLAM and AlereLAM tests. Results of FujiLAM tests were not communicated to the managing clinical team, but AlereLAM results were, if AlereLAM formed part of the local country guidelines of the study site (Malawi, South Africa, Tanzania, Thailand, Uganda, and Zambia).

Invalid FujiLAM or AlereLAM tests were repeated once. Further details of testing and operator training for both tests are included in the methods section of the online data supplement.

#### Reference standard testing

For reference standard testing, the specimens were processed using standardized protocols from centralized accredited laboratories of the different partner sites. Sputum, blood, and urine specimens were collected to ensure comprehensive reference standards.

Reference standard positives and negatives were defined as per Supplementary Table E2. Results from additional non-study specimens were captured for eMRS classification.

#### Post-hoc assessment of lot-to-lot variability

Variability across the six FujiLAM lots was assessed at the Research Institute of Tuberculosis of the Japan Anti-Tuberculosis Association (RIT-JATA, Tokyo, Japan). For this post-hoc analysis 181 urine samples were selected: 111 were FujiLAM positive but eMRS negative from this study (Supplementary Table E3) and an additional 70 were well-characterized samples from the FIND biobank of 50 microbiologically confirmed TB and 20 non-TB patients (see the procedures section in the online data supplement for more detail). All 181 urine samples were tested on each of the six FujiLAM lots and the AlereLAM test in singlicate. Each test was interpreted by two operators independently and in case of discordant results, the operators reinspected the test strip together to establish the final consensus result through mutual agreement. Operators were blinded to the initial result of the 181 samples. LAM concentration was further quantified using the ultrasensitive laboratory-based electrochemiluminescence LAM assay (EclLAM, Meso Scale Diagnostics, Rockville, MD, USA) employing the same antibody pair as the FujiLAM assay (12, 16).

#### Statistical analysis

A sample size of 233 confirmed TB patients across all sites was considered adequate to obtain an estimate of 60% (+/_□9%) overall FujiLAM sensitivity(11) with 95% Wilson’s Confidence Interval (CI), 80% power and 5% alpha.

Descriptive statistics were used to characterize participants. Index test sensitivity and specificity were determined using MRS, eMRS and CRS as reference standards. Overall sensitivity and specificity were calculated by pooling results from different sites. Results are presented with 95% CI based on Wilson’s score method.

Patients with invalid LAM-based test results and/or with all reference test results being contaminated or invalid were excluded from the respective analysis.

Data analysis was performed with R (version 4·1·2) based on a predefined statistical analysis plan and reported according to STARD guidelines (17). The statistical analysis plan is available upon request. The performance analysis by lot was done post-hoc.

Generalized linear mixed models (GLMM) were constructed post-hoc to investigate factors contributing to the variation in the agreement (match/mismatch) between the reference (eMRS) and FujiLAM using “lme4” package, “glmer” function, with a binomial error distribution (18). Age, sex, country, lot, visit (Day 1/Day 2), CD4 counts (log-transformed), urine colour, urine turbidity, and hospitalization setting (inpatient/outpatient) were included as fixed effects, patient ID was included as a random effect, and the test reader was included as a random effect nested within the country. Model summaries include fixed effect coefficients, standard errors, z-values and associated p-values, odds ratios, and their 95% CIs.

## Results

Across the study sites, 3528 PLHIV at risk of having pulmonary and/or extra-pulmonary TB were screened for eligibility. Of these, 1732 participants consented to participate in the study (Figure 1).

**Figure 1.**
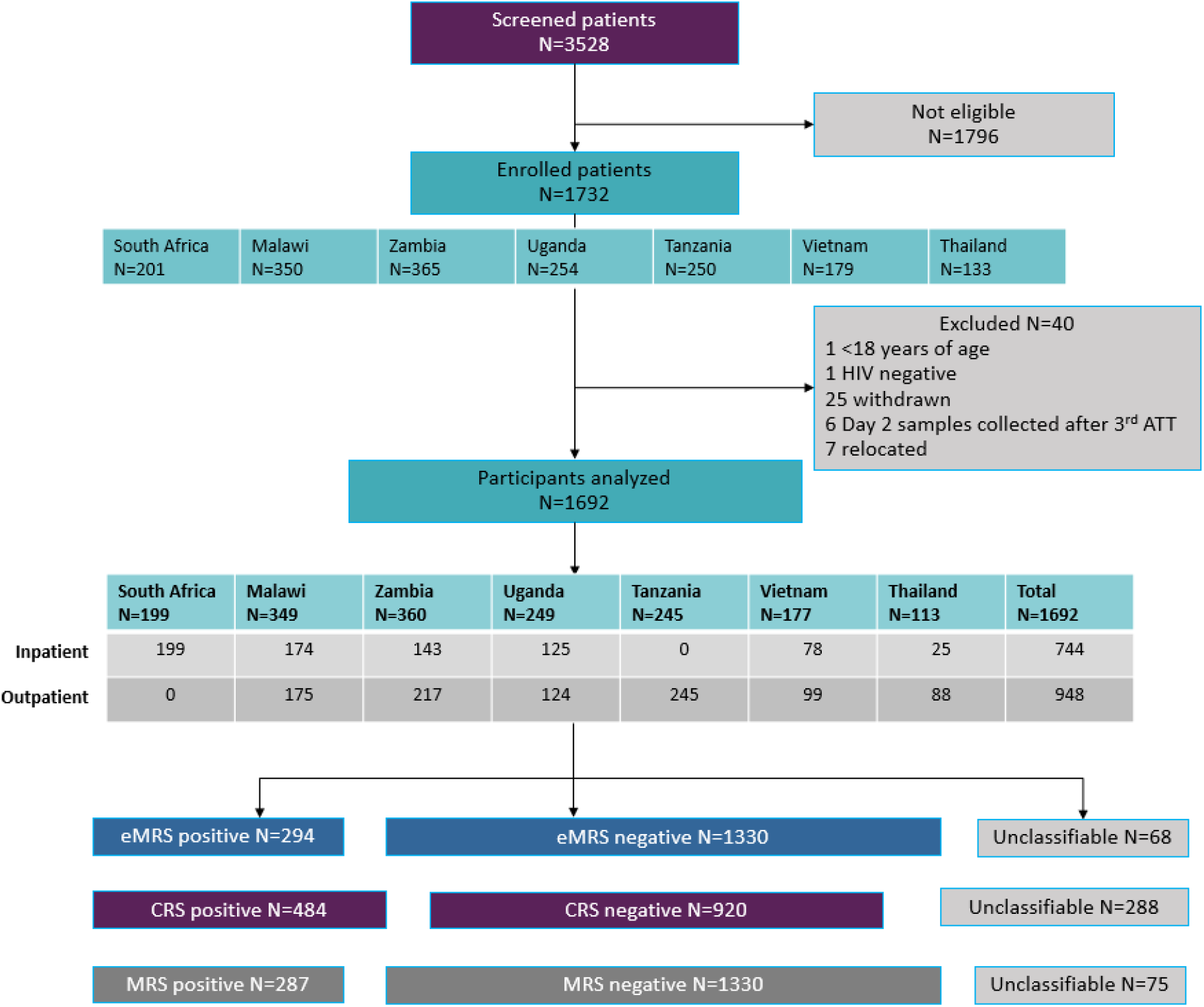
Study flow diagram. ATT, anti-TB therapy; CRS, composite reference standard; eMRS, extended microbiological reference standard; MRS, microbiological reference standard. Unclassifiable is neither reference standard positive nor reference standard negative. Reasons for non-eligibility of the 1796 persons screened but not enrolled were: being HIV negative, not interested to participate in the study, already on anti-TB treatment, on isoniazid preventive therapy, not willing to come back for follow-up visit, too weak or confused, refused to give blood.

A total of 1624 participants had results for all index and eMRS tests available and were included in the analysis. Table 1 shows their baseline demographic and clinical characteristics. The median age was 40 years (range: 18–82 years) and 52% were female. Overall, 26% had a history of prior TB treatment, 5% had a history of prior ART and 78% were on ART at the date of consent. Median CD4 count was 372 cells/µl. Of the 1624 participants, 294 (18.0%) were classified as positive for TB by eMRS and 1330 (82.0%) as negative for TB. Supplementary Table E4 shows the demographic and clinical characteristics of the study participants stratified by country.

**Table 1.**
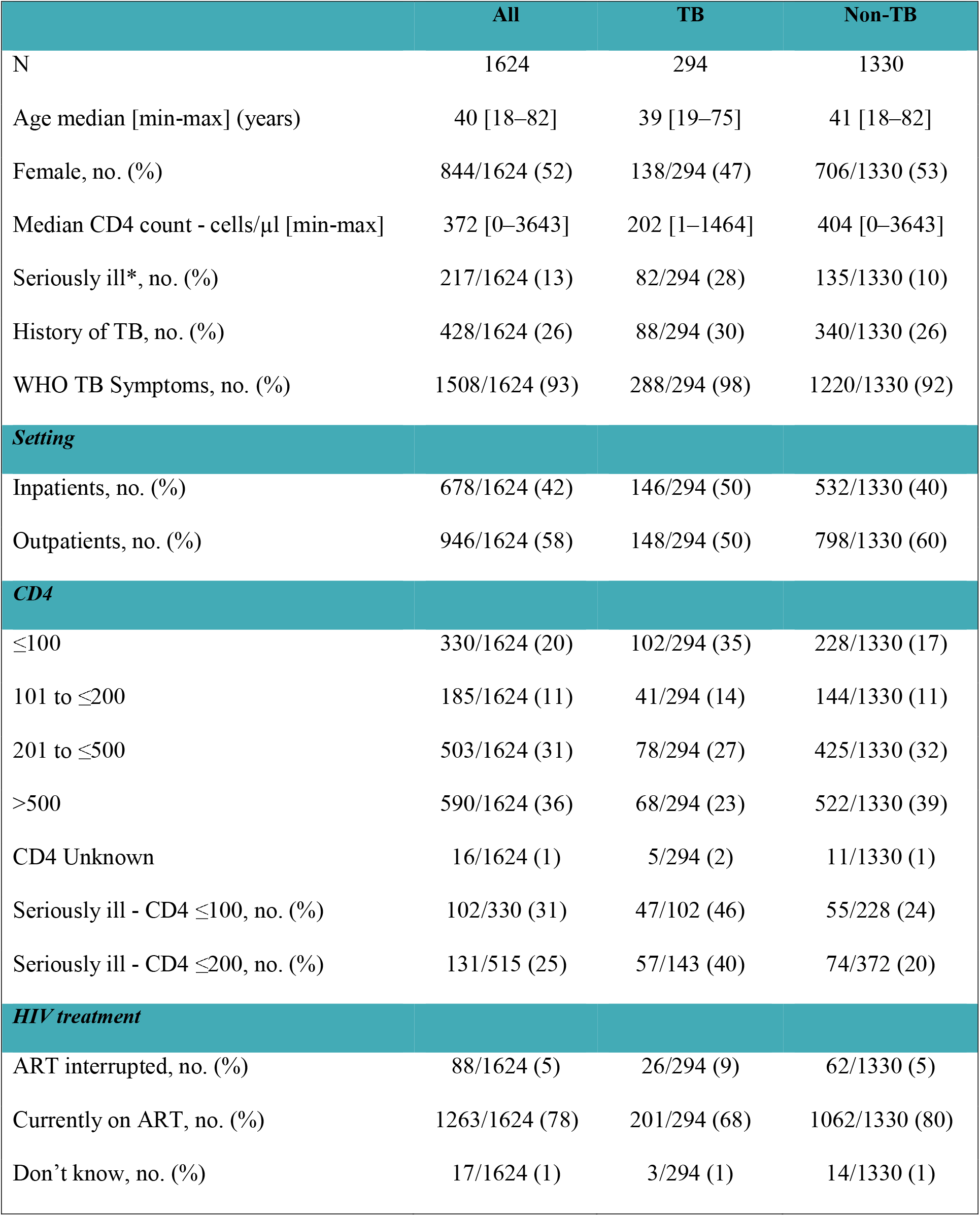
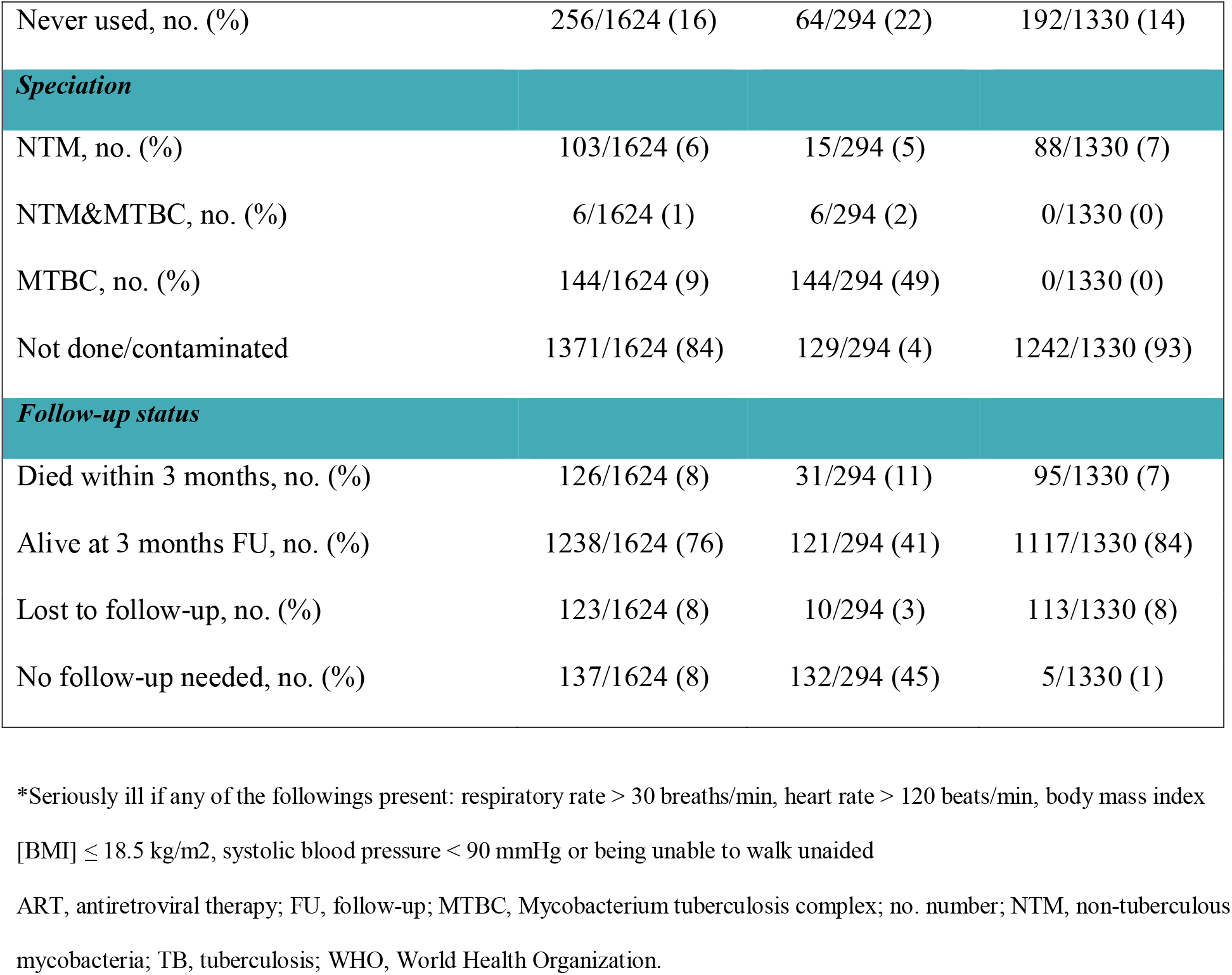
Demographic and clinical characteristics of study participants.

|  | All | TB | Non-TB |
| --- | --- | --- | --- |
| N | 1624 | 294 | 1330 |
| Age median [min-max] (years) | 40 [18–82] | 39 [19–75] | 41 [18–82] |
| Female, no. (%) | 844/1624 (52) | 138/294 (47) | 706/1330 (53) |
| Median CD4 count - cells/μl [min-max] | 372 [0–3643] | 202 [1–1464] | 404 [0–3643] |
| Seriously ill*, no. (%) | 217/1624 (13) | 82/294 (28) | 135/1330 (10) |
| History of TB, no. (%) | 428/1624 (26) | 88/294 (30) | 340/1330 (26) |
| WHO TB Symptoms, no. (%) | 1508/1624 (93) | 288/294 (98) | 1220/1330 (92) |
| **Setting** |  |  |  |
| Inpatients, no. (%) | 678/1624 (42) | 146/294 (50) | 532/1330 (40) |
| Outpatients, no. (%) | 946/1624 (58) | 148/294 (50) | 798/1330 (60) |
| **CD4** |  |  |  |
| $\leq 100$ | 330/1624 (20) | 102/294 (35) | 228/1330 (17) |
| 101 to $\leq 200$ | 185/1624 (11) | 41/294 (14) | 144/1330 (11) |
| 201 to $\leq 500$ | 503/1624 (31) | 78/294 (27) | 425/1330 (32) |
| $> 500$ | 590/1624 (36) | 68/294 (23) | 522/1330 (39) |
| CD4 Unknown | 16/1624 (1) | 5/294 (2) | 11/1330 (1) |
| Seriously ill - CD4 $\leq 100$, no. (%) | 102/330 (31) | 47/102 (46) | 55/228 (24) |
| Seriously ill - CD4 $\leq 200$, no. (%) | 131/515 (25) | 57/143 (40) | 74/372 (20) |
| **HIV treatment** |  |  |  |
| ART interrupted, no. (%) | 88/1624 (5) | 26/294 (9) | 62/1330 (5) |
| Currently on ART, no. (%) | 1263/1624 (78) | 201/294 (68) | 1062/1330 (80) |
| Don't know, no. (%) | 17/1624 (1) | 3/294 (1) | 14/1330 (1) |
| Never used, no. (%) | 256/1624 (16) | 64/294 (22) | 192/1330 (14) |
| ***Speciation*** |  |  |  |
| NTM, no. (%) | 103/1624 (6) | 15/294 (5) | 88/1330 (7) |
| NTM&MTBC, no. (%) | 6/1624 (1) | 6/294 (2) | 0/1330 (0) |
| MTBC, no. (%) | 144/1624 (9) | 144/294 (49) | 0/1330 (0) |
| Not done/contaminated | 1371/1624 (84) | 129/294 (4) | 1242/1330 (93) |
| ***Follow-up status*** |  |  |  |
| Died within 3 months, no. (%) | 126/1624 (8) | 31/294 (11) | 95/1330 (7) |
| Alive at 3 months FU, no. (%) | 1238/1624 (76) | 121/294 (41) | 1117/1330 (84) |
| Lost to follow-up, no. (%) | 123/1624 (8) | 10/294 (3) | 113/1330 (8) |
| No follow-up needed, no. (%) | 137/1624 (8) | 132/294 (45) | 5/1330 (1) |

\*Seriously ill if any of the followings present: respiratory rate > 30 breaths/min, heart rate > 120 beats/min, body mass index [BMI] ≤ 18.5 kg/m<sup>2</sup>, systolic blood pressure < 90 mmHg or being unable to walk unaided
ART, antiretroviral therapy; FU, follow-up; MTBC, Mycobacterium tuberculosis complex; no. number; NTM, non-tuberculous mycobacteria; TB, tuberculosis; WHO, World Health Organization.

### Diagnostic accuracy of FujiLAM

Overall sensitivity of FujiLAM against the eMRS on Day 1 was 54·8% (95% CI: 49·1–60·4), with an overall specificity of 85·1% (83·1–86·9) (Table 2). The Day 2 early morning sample had lower sensitivity and specificity estimates: 51·4% (45·6–57·1) and 81·7% (79·5–83·7), respectively (Supplementary Table E5). In comparison, overall sensitivity of AlereLAM against the eMRS on Day 1 was 30·5% (25·5–36·0), with an overall specificity of 90·7% (89·0–92·2). On Day 2 early morning urine samples, the sensitivity of AlereLAM was 28.4% (23·5–33·8) and specificity was 87·5% (85·6–89·2) (Supplementary Table E6).

**Table 2.** Sensitivity and specificity of Day 1 FujiLAM against the eMRS.

|  | N | TP | FP | FN | TN | Sensitivity [95%CI] | Specificity [95%CI] |
| --- | --- | --- | --- | --- | --- | --- | --- |
| All | 1615 | 160 | 197 | 132 | 1126 | 54.8 [49.1–60.4] | 85.1 [83.1–86.9] |
| ***CD4*** |  |  |  |  |  |  |  |
| ≤100 | 326 | 83 | 44 | 18 | 181 | 82.2 [73.6–88.4] | 80.4 [74.8–85.1] |
| 101 to ≤200 | 183 | 25 | 18 | 15 | 125 | 62.5 [47.0–75.8] | 87.4 [81.0–91.9] |
| 201 to ≤500 | 502 | 35 | 70 | 43 | 354 | 44.9 [34.3–55.9] | 83.5 [79.7–86.7] |
| >500 | 588 | 15 | 64 | 53 | 456 | 22.1 [13.9–33.3] | 87.7 [84.6–90.2] |
| Unknown | 16 | 2 | 1 | 3 | 10 | 40.0 [11.8–76.9] | 90.9 [62.3–98.4] |
| ***Setting*** |  |  |  |  |  |  |  |
| Inpatient | 671 | 100 | 63 | 44 | 464 | 69.4 [61.5–76.4] | 88.0 [85.0–90.5] |
| Outpatient | 944 | 60 | 134 | 88 | 662 | 40.5 [33.0–48.6] | 83.2 [80.4–85.6] |
| ***Country*** |  |  |  |  |  |  |  |
| South Africa | 144 | 41 | 22 | 15 | 66 | 73.2 [60.4–83.0] | 75.0 [65.0–82.9] |
| Malawi | 334 | 20 | 36 | 12 | 266 | 62.5 [45.2–77.1] | 88.1 [83.9–91.3] |
| Zambia | 358 | 33 | 56 | 20 | 249 | 62.3 [48.8–74.1] | 81.6 [76.9–85.6] |
| Uganda | 248 | 32 | 36 | 12 | 168 | 72.7 [58.1–83.7] | 82.4 [76.5–87.0] |
| Tanzania | 245 | 18 | 23 | 50 | 154 | 26.5 [17.4–38.0] | 87.0 [81.3–91.2] |
| Viet Nam | 176 | 11 | 5 | 22 | 138 | 33.3 [19.8–50.4] | 96.5 [92.1–98.5] |
| Thailand | 110 | 5 | 19 | 1 | 85 | 83.3 [43.6–97.0] | 81.7 [73.2–88.0] |

Subsequent sub-group analyses for Day 1 samples against eMRS, Day 2 results and estimates against MRS and CRS are reported in the Supplementary Tables E5, E7, and E9.

Stratified by CD4 count, FujiLAM sensitivity was 82·2% (73·6–88·4) and specificity was 80·4% (74·8–85·1) in participants with a CD4 count ≤100 cells/µl; sensitivity decreased at higher CD4 count strata, while specificity varied as shown in Table 2. In participants with CD4 201–500 cells/µl, sensitivity of FujiLAM was 44·9% (34·3–55·9) and specificity was 83·5% (79·7–86·7), while with CD4>500 cells/µl sensitivity was 22·1% (13·9–33·3) with 87·7% (84·6– 90·2) specificity (Table 2).

When stratified by setting, for inpatients, sensitivity of FujiLAM was 69·4% (61·5–76·4) and specificity was 88·0% (85·0–90·5). However, for outpatients, the tests showed reduced sensitivity and specificity at 40·5% (33·0–48·6) and 83·2% (80·4–85·6), respectively (Table 2).

Accuracy estimates varied considerably between countries, with sensitivity ranging from 26·5% (Tanzania), to 83·3% (Thailand), and specificity from 75·0% (South Africa) to 96·5% (Viet Nam, Table 3). Because the lot distribution was uneven between countries and could explain these differences (Supplementary Figure E2), in post-hoc analysis we calculated FujiLAM accuracy by lot. This identified substantial FujiLAM lot-to-lot variability, with certain lots delivering low specificity/high sensitivity and others delivering high specificity/low sensitivity (Figure 2).

**Figure 2.**
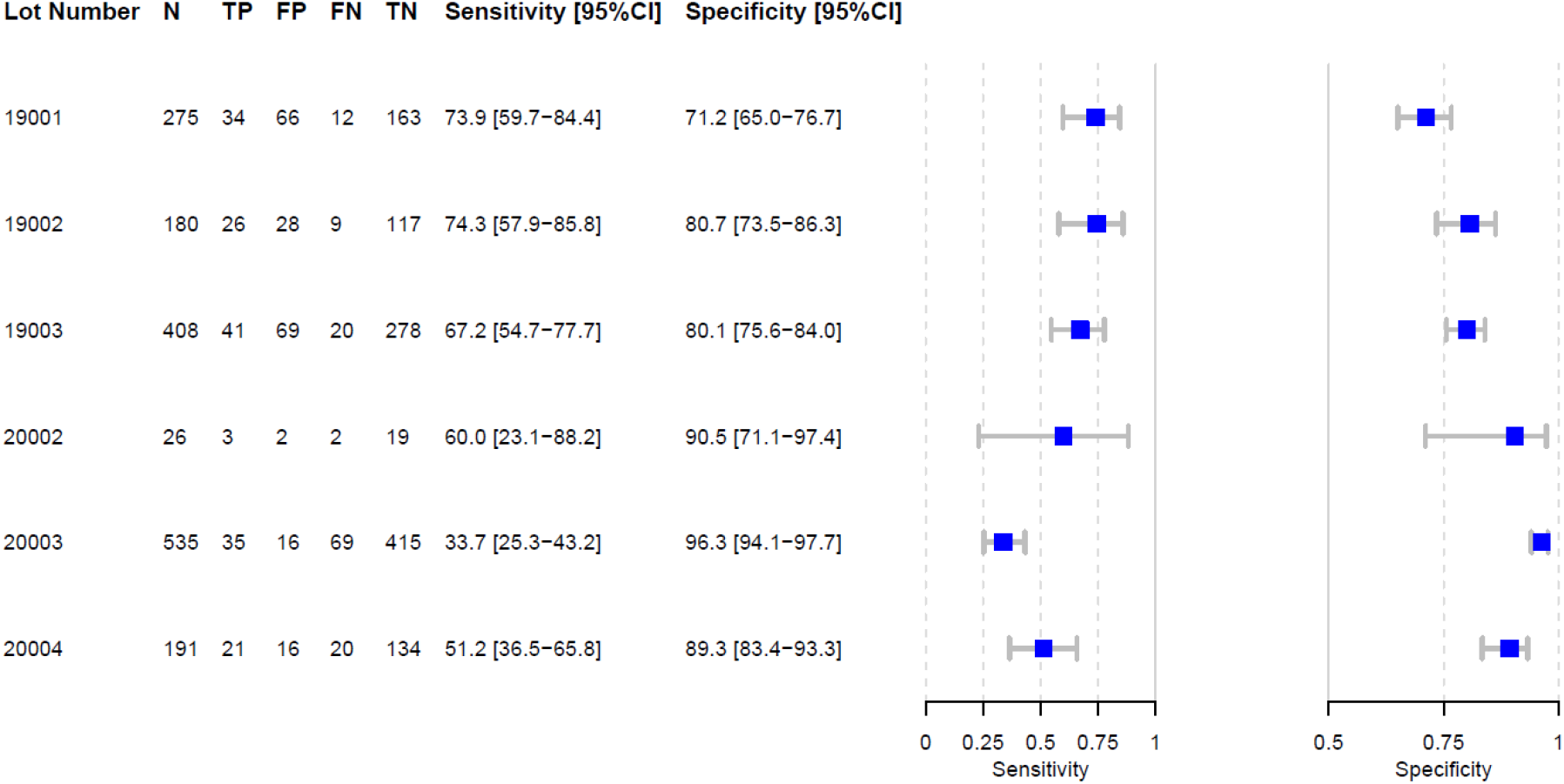
FujiLAM performance by lot. FN, false negative; FP, false positive; N, number; TN, true negative; TP true positive.

The diagnostic accuracy analysis of AlereLAM results on Day 1 and Day 2 against eMRS, CRS and MRS are reported in Supplementary Tables E6, E8 and E10, respectively. These analyses also reveal country-specific differences, however, to a lesser degree than for FujiLAM. Table E11 shows the number of additional microbiological tests per country considered for the eMRS, done by the routine clinical team.

### Additional post-hoc exploratory analysis

The regression model suggested that the factors significantly contributing to the variation between reference standard and FujiLAM were lot number (χ^2^_5_ =51·1 *p*=7·16×10^−10^), countries (χ^2^_6_=38·8, *p*=7·81×10^−7^) and visit days (χ^2^_1_=9·10, *p*=0·0025) (Supplementary Tables E12 and E13). However, because lots were not evenly distributed across countries, these factors may be interdependent, and the variation between different countries may be explained by the variation between lots or vice versa.

For eMRS positive patients (focusing on sensitivity), the only factor that remained significant was CD4 count (Supplementary Tables E14 and E15), where higher CD4 count was associated with a higher mismatch ratio (see Supplementary Figure E3). However, for eMRS negative patients (focusing on specificity), lot remained the most significant factor (χ^2^_5_=97·2, *p*<2·02×10^−19^), thus explaining the variation in agreement between the reference standard and the FujiLAM test (Supplementary Tables E16 and E17).

To verify the impact of lots on performance, we analysed 111 FujiLAM-positive, eMRS-negative urine specimens from the study on all six lots used in the study and with AlereLAM. As shown in Figure 3 and Supplementary Table E18, FujiLAM positivity rates varied from 12/111 (13%) to 86/111 (77%) between lots. In addition, we quantified the concentration of LAM in 110 of the 111 samples using EclLAM (one sample was not available for EclLAM testing due to insufficient volume). A total of 14 samples had measurable LAM concentration (>11 pg/mL); of these, 12 were concordant positive on all six FujiLAM lots tested, of which three were further classified as CRS positive, eight as CRS negative and three as CRS unclassifiable (patient passed away). Twenty-one of the 111 samples tested negative on all six FujiLAM lots.

**Figure 3.**
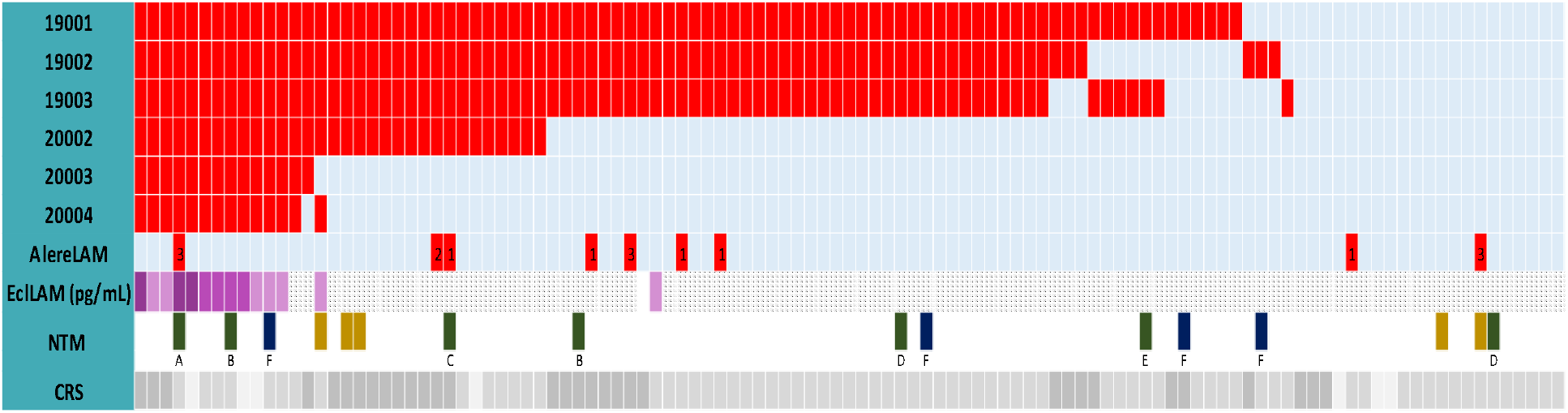
Exploratory comparison of FujiLAM positivity rates in 111 eMRS negative, FujiLAM positive samples from the study. eMRS, extended microbiological reference standard; NTM, nontuberculous mycobacteria; CRS, composite reference standard; Red cells indicate positive result and light blue cells indicate negative result on FujiLAM or AlereLAM. For AlereLAM positive results, the numbers further indicate line grade intensity (1–3). LAM concentration measured with EclLAM is illustrated on the purple scale from darkest to lightest: >200 pg/mL, 51–200 pg/mL, 11–50 pg/mL; diagonal stipe pattern indicates <limit of detection (11 pg/mL); Dark yellow – Full NTM speciation not done; green - slow growing mycobacteria; dark blue-fast growing mycobacteria; A-*M. simiae*; B-*M. intracellulare*; C-*M. avium*, D-*M. scrofulaceum*; D-*M. gordonae*; F-*M. fortuitum;* Dark grey-CRS positive, mid grey-CRF neg, light grey-unclassifiable.

Based on the data available, there is no clear indication that false positivity correlates with the presence of non-tuberculous mycobacteria (NTM) infection measured in sputum samples. However, four of the eight CRS-negative, FujiLAM-positives had LAM levels above the detection limit of the EclLAM assay and had NTM detected, but notably from a non-sterile sputum sample only; two were slow-growing, one was fast-growing, and one was non-specified NTM.

We furthermore determined FujiLAM (six lots) and AlereLAM positivity rates in a series of well-characterized biobank specimens from 50 patients with microbiologically confirmed TB and 20 patients with negative microbiological test results (Supplementary Table E19). This experiment confirmed that high positivity rates were associated with certain lots.

## Discussion

This is the first prospective, multi-centre diagnostic accuracy study of the FujiLAM test in PLHIV, conducted among inpatient and outpatient settings in seven countries across sub-Saharan Africa and Asia. We observed considerably higher sensitivity of the FujiLAM test than AlereLAM (a difference of 24%); however, its specificity was lower than expected from previously published studies (11, 14, 19). Furthermore, we observed a large variability in FujiLAM sensitivity and specificity between countries, which was attributed to variable performance between FujiLAM lots, limiting our interpretation of study findings.

This is the first study reporting variable performance with different FujiLAM lots. Earlier studies mainly used single lots, none of which were used in this study (Supplementary Table E20). Reported sensitivity and specificity results have been consistent across past studies. A recent meta-analysis, including five adult cohorts of PLHIV from three countries using three different lots (98002, 98004, 98006), found no inconsistency in diagnostic accuracy (14). In another study from Nigeria, sub-group analysis of FujiLAM performance by HIV status, using lot 20001, showed 93·3% specificity (20), comparable to previous studies and to specificity values of lots 20002, 20003 and 20004 used in this study.

Assessment of lot variability was not considered in the conceptualization of the study; however, the large study size allowed for exploratory analysis of factors that could explain the unexpected clinical variability. Post-hoc re-analysis of 111/197 samples deemed false positive in the clinical study, and an additional 70 representative samples from the FIND biobank confirmed a significant difference in positivity between the six lots used in the study. EclLAM, a quantitative research assay employing the same antibodies as FujiLAM, only detected LAM in 14 of the 110 eMRS-negative FujiLAM-positive samples from the study. Twenty-one of the 111 samples tested negative on all six FujiLAM lots, all of these had LAM concentration measured by EclLAM below the limit of detection.

Overall, we found higher specificity of AlereLAM compared with FujiLAM (a difference of 5.6%), and we observed some variability of AlereLAM specificity at country level (81%–95%), although much less compared with FujiLAM (75–97%). Previously published studies in PLHIV found consistently lower specificity of FujiLAM compared with AlereLAM for those with CD4

<200 cells/°L (a difference of 13.1% for CD4 0–100; and 1.7% for CD4 100–200) (11, 14, 19). This was not observed in PLHIV with high CD4 cell counts or in patients without HIV(12) and has been interpreted in parts to an effect driven by the imperfect reference standard for TB diagnosis, which disproportionally affects more sensitive tests and results in lower specificity (11, 14, 21, 22).

As the FujiLAM test is visually read and interpreted, it is not possible to adjust interpretation of specific lots, as with most lateral flow tests using a reference card (such as AlereLAM)(23) or computerized reader. Current and future LAM tests may benefit from a reading device, which could improve consistency and remove reader subjectivity, particularly for bands close to the low cut-off in the pg/ml range, which is required for LAM tests to reach sufficient sensitivity (16). A reader device with connectivity can further enable automated linkage of test results to care, as well as improved surveillance.

The observed FujiLAM lot variability can impact patient management. Translating the findings to a setting with 10% prevalence of TB among patients presenting for care, the most extreme performing lots, 19001 (sensitivity: 73·9 [59·7–84·4]; specificity: 71·2 [65·0–76·7]) and 20003 (sensitivity: 33·7 [25·3–43·2]; specificity: 96·3 [94·1–97·7]) would render large differences in test outcomes. For each 1000 tested individuals, lot 19001 would identify 74 (95% CI: 60–84) true positive and 26 (95% CI: 16 to 40) false negatives, whereas lot 20003 would identify 34 (26–43) true positive and 66 (57–74%) false negatives. More worryingly, lot 19001 would identify 259 (210–315) false positives, whereas lot 20003 would only identify 33 (21–53) false positives (Supplementary Table E21 and E22). This variability is unacceptably high for clinical management of patients.

Several ongoing studies are evaluating the accuracy of FujiLAM and specific lot analyses will be important to verify the findings from this study. Altogether, this study underlines the importance of conducting manufacturer-independent evaluations of new diagnostic tests. When designing a diagnostic accuracy study, it is critical to include at least two lots evenly distributed across the clinical sites and systematic quality control using external reference material. To our knowledge, there are currently no available quality assessment panels for a LAM-based test.

In conclusion, this large multi-country clinical trial of the diagnostic accuracy of the FujiLAM test observed higher FujiLAM sensitivity in PLHIV with low CD4 cell counts and in inpatients, in accordance with previous studies. However, specificity was lower than expected, and accuracy estimates were variable and associated with specific FujiLAM lots, as confirmed through additional post-hoc testing and analysis. The lot variability issue with the FujiLAM test is a major setback in the quest towards a POC non-sputum-based TB test. Although the results obtained using the current version of the FujiLAM test are too variable for clinical decision-making, a new version of the test (work already undergoing by the manufacturer) could improve POC testing for TB diagnosis in PLHIV. Despite these challenges and unexpected observations, it is important to emphasize the promise that the LAM biomarker and LAM tests hold for TB testing.

## Data Availability

Individual, de-identified participant data will be shared, including data dictionaries. Other documents that have been made available include the study protocol and statistical analysis plan. Templates of the informed consent forms may be shared upon request. The data will be available immediately following publication with no end date. The data will be shared with anyone who wishes to access the data. The data will be available for any purpose of analyses. For data, please contact the corresponding author.

## Acknowledgements

The authors would like to thank the study participants and their families for generously volunteering to participate in this study, as well as the study sites for their time and effort in conducting the study and assisting with the analysis of the operational data.

We also thank Pamela Nabeta, Samuel Schumacher for their scientific input in study design; Agnes Malobela, Geetanjali Kataria, Sunita Singh and Divya Soni for their study monitoring support. We thank Rene Goliath for study support at the UCT site, Marriott Nliwasa, Elizabeth L Corbett and Ailva O’Reilly at the Malawi site. Medical writing support was provided by Talya Underwood, MPhil, of Anthos Communications Ltd, UK, funded by FIND, according to Good Publication Practice guidelines.

