## Supplementary material for "Multicentre accuracy trial of FUJIFILM SILVAMP TB LAM test in people with HIV reveals lot variability": Online data supplement

**Contents**

Supplementary methods: page 3

FujiLAM proficiency tool: page 4

AlereLAM proficiency tool: page 12

Figures and tables: page 18

Supplementary references: page 45

**Supplementary methods**

***Procedures***

The 70 urine samples selected from the FIND biobank were representatives of both microbiologically confirmed TB and non-TB patients. All 70 samples had their LAM concentration previously determined by the EclLAM assay, thus they also served as control material for the post-hoc lot-to-lot assessment. A total of 47 of the 50 microbiologically confirmed TB samples had both sputum culture and smear positive results and three of the 50 TB positives were confirmed by Xpert MTB/RIF. Their selection mainly centred around the 10–50 pg/mL LAM concentration range, which includes the limit of detection (LoD) of the FujiLAM test (~ 30 pg/mL) as lot-to-lot variability is more likely to impact the threshold for LAM detection. The remaining 20 samples were TB-negative (as per MRS) with undetectable LAM concentration by the EclLAM assay (below the LoD of 11pg/mL), thus ideally should not yield a signal with the FujiLAM test.

***FujiLAM testing***

FujiLAM testing was done as per manufacturer’s instructions. Briefly, urine was added to the reagent tube up to the indicator line (approximately 200 µl), mixed, and incubated for 40 minutes at ambient temperature. After mixing again, two drops of urine/reagent are added to the test strip. Following this, a button is pressed immediately to release a reducing agent for silver amplification. After the colour indicator mark turns orange (approximately 10 minutes), the next button is pressed. The result is then read within 10 minutes. The FujiLAM assay does not use a reference scale card and any line seen on the test is considered positive.

Operators were trained prior to the study start and their competency was assessed using a proficiency testing tool (see page 3) composed of the following four sections:

1. FujiLAM test run on a blinded mock urine sample observed by a moderator
2. Appraisal of the moderator
3. Questionnaire to assess operator’s understanding of the FujiLAM test procedure
4. Test result interpretation using photographs of FujiLAM results.

***AlereLAM testing***

AlereLAM testing was done as per manufacturer’s instructions. Operators were trained prior to the study start and their competency was assessed using a proficiency testing tool (see page 11) composed of the following 4 sections:

1. AlereLAM test run on a blinded mock urine sample observed by a moderator
2. Appraisal of the moderator
3. Questionnaire to assess operator’s understanding of the AlereLAM test procedure
4. Test result interpretation using photographs of AlereLAM results.

Both FujiLAM and AlereLAM tests were done within 2 hours of sample collection at room temperature.

***Statistical analysis***

Variables such as age, sex, country, visit day, CD4 cell count and hospitalization were included as they have previously been shown to be related to the accuracy of diagnostic tests. Variables such as lot and reader were included to assess whether the variability was related to manufacturing or training procedures/readers’ performance. Additionally, urine characteristics were included to investigate whether the presence of particles or blood in the urine affected test outcome.

To investigate whether the sensitivity and specificity of FujiLAM differed significantly for the same reasons, we fitted GLMMs in TB positive and negative patients (per eMRS reference) separately.

The relationship between the CD4 counts and FujiLAM/eMRS result mismatch was further investigated by a linear regression, using the “lm” function in the base package. CD4 counts were categorized based on previous thresholds for visual representation. The linear model was constructed using the median CD4 value of each category.

**FujiLAM proficiency testing tool**

**FUJIFILM SILVAMP TB LAM (FujiLAM) – PROFICIENCY TESTING TOOL**

**Intended use**

This tool is intended to be used by the moderator to assess the minimum training needs and the resulting proficiency of health care workers who are intended to be end users of the FUJIFILM SILVAMP TB LAM (FujiLAM) test. This proficiency assessment will be carried out after initial training. Once operators have processed at least two FujiLAM tests during the training, each operator will be asked to independently perform one complete FujiLAM test runs using blinded mock urine sample. The moderator will observe, without intervening or correcting mistakes.

The proficiency testing comprise of the following parts:

A. **Observed hands-on FujiLAM test run**. Each operator will be provided with one blinded mock urine sample and asked to perform the FujiLAM test. The moderator will observe the test run and complete a proficiency checklist to assess adherence to the critical steps according to the FujiLAM Quick Reference Guide/Instructions for Use (IFU)/video to ensure reliable test results.
B. **Appraisal**. At the end, the moderator will estimate the level of confidence shown by the operator while performing the FujiLAM tests.
C. **Questionnaire**. 10 questions will be asked orally by the moderator after the test run to assess the level of understanding of the FujiLAM test and the operators’ ability in coping with problems that might arise during the procedure.
D. **Test result interpretation**. Each operator will be provided with photographs of possible test results including positive, negative and invalid. Although the photographs are not the same as test results in real, this will allow the assessment of results interpretation of different band intensities.

**Performance targets**

The training will be considered successful if the following performance targets are met:

- Individual scores for Part A, C and D ≥ 22 (80%), ≥8/10 (80%) and ≥9 (90%), respectively
- **AND** an overall appraisal of the operator’s confidence to perform FujiLAM of ≥ 4 (on a scale of 1-5)

If performance targets are not met after initial training, the operator will undergo additional training on specific topics, and be reassessed for proficiency until targets are met.

**Materials needed**:

| **FujiLAM kit (materials included)** | **Materials not provided** |
| --- | --- |
| Pipette | Urine collection cup |
| Tube Rack | Gloves |
| Nozzle | Permanent marker |
| Quick Reference Guide | Biohazard waste bin |
| Instructions For Use | Artificial urine samples ( 10 ml of 5 ng/ml LAM in buffer as positive control and 10 ml of sterile water as negative control) |
| Test pouch (test cartridge & reagent tube) |  |

A. **Observed hands-on FujiLAM test run – Checklist**

**Instructions:**

- Prepare the workspace.
- Process 1 sample according to the FujiLAM Quick Reference Guide/Instructions for Use.
- Two moderators will be present: One of them will video record the session and the other will complete the checklist below.
- The operator has to perform the tasks outlined in the checklist correctly. If not, the answer should be “NO” and an explanation should be added on the last column.
- For each correctly performed item, the operator will obtain 1 point.

NAME OF OPERATOR: __________________________	DATE OF COMPLETION: _________________

| **Procedure** | **Step** | **Assessment of sample (If NO add comment)** | | **Comment** |
| --- | --- | --- | --- | --- |
| **FujiLAM preparation** | 1. Did the operator collect all necessary materials (the kit and additional materials needed including urine sample) as outlined in the instructions? | ❑YES | ❑NO |  |
|  | 2. Did the operator open the cardboard box correctly? | ❑YES | ❑NO |  |
|  | 3. Did the operator put on appropriate protective equipment (gloves, lab coat)? | ❑YES | ❑NO |  |
|  | 4. Did the operator check the expiry date on the test pouch? | ❑YES | ❑NO |  |
|  | 5. Did the operator open the test pouch | ❑YES | ❑NO |  |
|  | 6. Did the operator remove both test cartridge and reagent tube from the pouch? | ❑YES | ❑NO |  |
|  | 7. Did the operator write clearly and at the appropriate place the patient ID on the test cartridge and reagent tube? | ❑YES | ❑NO |  |
| **FujiLAM sample preparation** | 8. Did the operator visually inspect the reagent tube for presence of a pad? | ❑YES | ❑NO |  |
|  | 9. Did the operator remove the seal of the reagent tube? | ❑YES | ❑NO |  |
|  | 10. Did the operator open the urine container at a time and close lid afterwards? | ❑YES | ❑NO |  |
|  | 11. Did the operator transfer urine to the correct tube? | ❑YES | ❑NO |  |
|  | 12. Was the correct volume of urine transferred to the reagent tube (up to indicator line)? | ❑YES | ❑NO |  |
|  | 13. Did the operator correctly attach the nozzle to the tube? | ❑YES | ❑NO |  |
|  | 14. Was the tube mixed gently 10 times, without inverting the tube? | ❑YES | ❑NO |  |
|  | 15. Did the operator incubate the sample for 40-50 minutes at room temperature? | ❑YES | ❑NO |  |
|  | 16. Was the tube, after the incubation, mixed gently again 10 times without inverting the tube? | ❑YES | ❑NO |  |
| **FujiLAM test procedure** | 17. Did the operator hold the reagent tube at 90 degrees to the test device while adding the drops? | ❑YES | ❑NO |  |
|  | 18. Did the operator hold the tube at about 1 cm height while adding the drops? | ❑YES | ❑NO |  |
|  | 19. Was the correct amount of the sample mixture (2 drops) added to the well (1) on the test cartridge? | ❑YES | ❑NO |  |
|  | 20. Was button (2) pushed immediately and completely by the operator until it became dented? | ❑YES | ❑NO |  |
|  | 21. Did the operator wait until the “Go Next” mark turned orange? | ❑YES | ❑NO |  |
|  | 22. Did the operator proceed to the next step within 30 mins? | ❑YES | ❑NO |  |
|  | 23. Was button (3) pushed completely by the operator until it became dented? | ❑YES | ❑NO |  |
| **FujiLAM interpretation of results** | 24. Was the result read within 1-10 minutes after the control line appeared? | ❑YES | ❑NO |  |
|  | 25. Was the operator able to correctly interpret the test result? | ❑YES | ❑NO |  |
|  | 26. Was the operator able to correctly record the test results and any comments on the FujiLAM result form? | ❑YES | ❑NO |  |
|  | 27. Did the operator dispose all urine specimens and assay materials in the appropriate biohazard waste bin? | ❑YES | ❑NO |  |
|  | 28. Did the operator disinfect the bench after use? | ❑YES | ❑NO |  |
| **PART A** | **Score / Number of correct items** | **/ 28** | | …………… % |

NAME OF MODERATOR: __________________________ DATE REVIEWED:_______________________

B. **Appraisal**

- The moderator that supervises the assessment will estimate the level of confidence is shown by the operator while performing the FujiLAM test.

NAME OF OPERATOR: __________________________	DATE OF COMPLETION: _________________

| **APPRAISAL** | How would you evaluate the level of confidence shown by the operator while performing FujiLAM?  ❑1 (not confident) ❑2 ❑3 ❑4 ❑5 (very confident)  Comments: |
| --- | --- |

NAME OF MODERATOR: __________________________ DATE REVIEWED:_______________________

C. **Questionnaire**

**Instructions:**

- The moderator will ask 10 out of the 29 following questions to each operator (individually) in the context of the FujiLAM proficiency run.
- For each correct item, the operator will obtain 1 point.

NAME OF OPERATOR: __________________________	DATE OF COMPLETION:_________________

| **FujiLAM Questions** | | **Answered correctly** | | **If NO add comment** |
| --- | --- | --- | --- | --- |
| 1. What is the intended use of FujiLAM? | | ❑YES | ❑NO |  |
| 2. What is the storage temperature of the kit? | | ❑YES | ❑NO |  |
| 3. Can you describe all the components that are included in the FujiLAM kit? | | ❑YES | ❑NO |  |
| 4. What kind of specimen is required for the FujiLAM test? | | ❑YES | ❑NO |  |
| 5. What do you do if the test device was dropped prior to testing? | | ❑YES | ❑NO |  |
| 6. Which lighting condition should be avoided while using the cartridge? | | ❑YES | ❑NO |  |
| 7. What would you do if the pouch of the device was not opened right before testing but well in advance i.e. contents exposed to ambient conditions? | | ❑YES | ❑NO |  |
| 8. Which two things do you need to check (visually) before carrying out the FujiLAM test? | | ❑YES | ❑NO |  |
| 9. What would you do if the expiry date of the test has passed? | | ❑YES | ❑NO |  |
| 10. What would you do if you realize the device is broken? | | ❑YES | ❑NO |  |
| 11. What would you do if you do not observe a pad in the reagent tube? | | ❑YES | ❑NO |  |
| 12. What would you do if the amount of urine that you transferred to the reagent tube is above the indicator line? | | ❑YES | ❑NO |  |
| 13. How critical is the urine volume added to the reagent tube? | | ❑YES | ❑NO |  |
| 14. What precaution do you need to take when mixing the reagent tube after adding urine? | | ❑YES | ❑NO |  |
| 15. What is the minimum time the urine has to be incubated in the reagent tube? | | ❑YES | ❑NO |  |
| 16. What is the maximum time you can let the reagent tube be incubated? | | ❑YES | ❑NO |  |
| 17. What would you do if you dropped the reagent tube containing the sample mixture and it splashed on your hands, face and/or eyes? | | ❑YES | ❑NO |  |
| 18. What would you do if you dropped the reagent tube containing the sample mixture and it splashed on the FujiLAM cartridge? | | ❑YES | ❑NO |  |
| 19. How many drops of the sample mixture should be transferred to the FujiLAM cartridge? | | ❑YES | ❑NO |  |
| 20. What would you do if you observe bubbles in the sample well (1)? | | ❑YES | ❑NO |  |
| 21. What should you do straight after transferring 2 drops of the sample mixture to the FujiLAM cartridge? | | ❑YES | ❑NO |  |
| 22. What would you do, if you pressed button (2) and then realized that you added only 1 drop of sample mixture? | | ❑YES | ❑NO |  |
| 23. How long should you wait after pressing button (2)? | | ❑YES | ❑NO |  |
| 24. What would you do if the ‘Go Next’ mark does not appear after 30 mins? | | ❑YES | ❑NO |  |
| 25. What is the next step after the ‘Go Next’ mark turns orange? | | ❑YES | ❑NO |  |
| 26. What would you do if, you realized that you read the orange ‘Go Next’ mark more than 30 minutes after you pressed button (2)? | | ❑YES | ❑NO |  |
| 27. What would you do if the control band does not appear after approximately 1 minute? | | ❑YES | ❑NO |  |
| 28. What is the maximum time within which you need to interpret the test results? | | ❑YES | ❑NO |  |
| 29. How should you dispose of the FujiLAM cartridge? | | ❑YES | ❑NO |  |
| **PART C** | **Score / Number of correct questions** | **/ 10** | | …………… % |

NAME OF MODERATOR: __________________________	DATE REVIEWED:_________________

D. **Test result interpretation**

**Instructions:**

- The moderator will provide a form containing 10 photographs of possible test results.
- The operator has to interpret the result for each case.
- For each correct item, the operator will obtain 1 point.

NAME OF OPERATOR: __________________________	DATE OF COMPLETION:_________________

|  | **Test result example** | **Result interpretation** | | | **Moderator’s comment** |
| --- | --- | --- | --- | --- | --- |
| 1 | 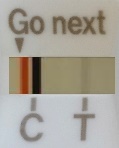 | ❑Positive | ❑Negative | ❑Invalid |  |
| 2 | 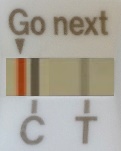 | ❑Positive | ❑Negative | ❑Invalid |  |
| 3 | 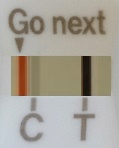 | ❑Positive | ❑Negative | ❑Invalid |  |
| 4 | 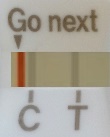 | ❑Positive | ❑Negative | ❑Invalid |  |
| 5 | 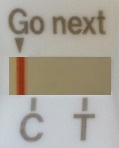 | ❑Positive | ❑Negative | ❑Invalid |  |
| 6 | 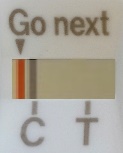 | ❑Positive | ❑Negative | ❑Invalid |  |
| 7 | 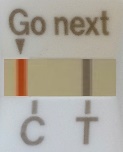 | ❑Positive | ❑Negative | ❑Invalid |  |
| 8 | 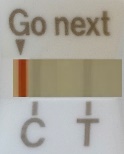 | ❑Positive | ❑Negative | ❑Invalid |  |
| 9 | 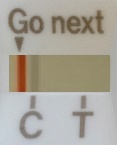 | ❑Positive | ❑Negative | ❑Invalid |  |
| 10 | 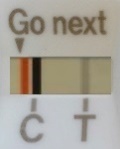 | ❑Positive | ❑Negative | ❑Invalid |  |
| **PART D** | | **Score / Number of correct items** | | **/ 10** | …………… % |

E. **Conclusion**

| **Performance targets met?** |  |  | **If NO add comment** |
| --- | --- | --- | --- |
| Score Part A: ≥80%? | ❑YES | ❑NO |  |
| Score Part B: Appraisal ≥4? | ❑YES | ❑NO |  |
| Score Part C: ≥80%? | ❑YES | ❑NO |  |
| Score Part D: ≥90%? | ❑YES | ❑NO |  |
| **Conclusion: Operator passed proficiency test** | **❑YES**^#^ | **❑NO** |  |

^#^Operator can only pass the proficiency test, when score for individual Parts A, B, C and D were ALL met.

NAME OF MODERATOR: __________________________	DATE ASSESSED:_________________

**AlereLAM proficiency testing tool**

**ALERE DETERMINE™ TB LAM (AlereLAM) – PROFICIENCY TESTING TOOL**

**Intended use**

This tool is intended to be used by the moderator to assess the minimum training needs and the resulting proficiency of health care workers who are intended to be end users of the ALERE DETERMINE™ TB LAM (AlereLAM) test. This proficiency assessment will be carried out after initial training. Once operators have processed at least two AlereLAM tests during the training, each operator will be asked to independently perform one complete AlereLAM test run using blinded mock urine sample. The moderator will observe, without intervening or correcting mistakes.

The proficiency testing comprise of the following parts:

E. **Observed hands-on AlereLAM test run**. Each operator will be provided with one blinded mock urine sample and asked to perform the AlereLAM test. The moderator will observe the test run and complete a proficiency checklist to assess adherence to the critical steps according to the AlereLAM Quick Reference Guide/Instructions for Use (IFU) to ensure reliable test results.
F. **Appraisal**. At the end, the moderator will estimate the level of confidence shown by the operator while performing the AlereLAM tests.
G. **Questionnaire**. 5 questions out of the 13 will be asked orally by the moderator after the test run to assess the level of understanding of the AlereLAM test and the operators’ ability in coping with problems that might arise during the procedure.
H. **Test result interpretation**. Each operator will be provided with photographs of possible test results including positive, negative and invalid. Although the photographs are not the same as test results in real, this will allow the assessment of results interpretation of different band intensities. Use the reference card for the interpretation.

**Performance targets**

The training will be considered successful if the following performance targets are met:

- Individual scores for Part A, C and D ≥ 22 (80%), ≥ 4 (80%) and ≥9 (90%), respectively
- **AND** an overall appraisal of the operator’s confidence to perform AlereLAM of ≥ 4 (on a scale of 1-5)

If performance targets are not met after initial training, the operator will undergo additional training on specific topics, and be reassessed for proficiency until targets are met.

**Materials needed**:

| **AlereLAM kit (materials included)** | **Materials not provided** |
| --- | --- |
| Instructions For Use | Urine collection cup |
| Test | Gloves |
| Reference Card | Permanent marker |
|  | Pipette |
|  | Biohazard waste bin |
|  | Artificial urine samples ( 10 ml of 5 ng/ml LAM in buffer as positive control and 10 ml of sterile water as negative control) |

F. **Observed hands-on FujiLAM test run – Checklist**

**Instructions:**

- Prepare the workspace.
- Process 1 sample according to the AlereLAM Instructions for Use.
- The moderator will complete the checklist below.
- The operator has to perform the tasks outlined in the checklist correctly. If not, the answer should be “NO” and an explanation should be added on the last column.
- For each correctly performed item, the operator will obtain 1 point.

NAME OF OPERATOR: __________________________	DATE OF COMPLETION: _________________

| **Procedure** | **Step** | **Assessment of sample (If NO add comment)** | | **Comment** |
| --- | --- | --- | --- | --- |
| **AlereLAM preparation** | 29. Did the operator collect all necessary materials (the kit and additional materials needed including urine sample) as outlined in the instructions? | ❑YES | ❑NO |  |
|  | 30. Did the operator open the aluminium pouch correctly? | ❑YES | ❑NO |  |
|  | 31. Did the operator put on appropriate protective equipment (gloves, lab coat)? | ❑YES | ❑NO |  |
|  | 32. Did the operator check the expiry date on the test pouch? | ❑YES | ❑NO |  |
|  | 33. Did the operator open the test tearing at the perforation? | ❑YES | ❑NO |  |
|  | 34. Did the operator remove the protecting foil cover completely? | ❑YES | ❑NO |  |
|  | 35. Did the operator write clearly and at the appropriate place the patient ID on the test strip? | ❑YES | ❑NO |  |
| **AlereLAM test procedure** | 36. Did the operator transfer 60 ul urine to the correct position of the test strip? | ❑YES | ❑NO |  |
|  | 37. Did the operator incubate the test in horizontal position? | ❑YES | ❑NO |  |
|  | 38. Did the operator wait 25 minutes before interpreting the results? | ❑YES | ❑NO |  |
|  | 39. Did the operator interpret the results within 35 minutes? | ❑YES | ❑NO |  |

| **AlereLAM interpretation of results** | 40. Was the operator able to correctly interpret the test result? | ❑YES | ❑NO |
| --- | --- | --- | --- |
|  | 41. Was the operator able to correctly record the test results and any comments on the AlereLAM result form? | ❑YES | ❑NO |
|  | 42. Did the operator dispose all urine specimens and assay materials in the appropriate biohazard waste bin? | ❑YES | ❑NO |
|  | 43. Did the operator disinfect the bench after use? | ❑YES | ❑NO |
| **PART A** | **Score / Number of correct items** | **/ 15** | …………… % |

NAME OF MODERATOR: __________________________ DATE REVIEWED:_______________________

G. **Appraisal**

- The moderator that supervises the assessment will estimate the level of confidence is shown by the operator while performing the AlereLAM test.

NAME OF OPERATOR: __________________________	DATE OF COMPLETION: _________________

| **APPRAISAL** | How would you evaluate the level of confidence shown by the operator while performing AlereLAM?  ❑1 (not confident) ❑2 ❑3 ❑4 ❑5 (very confident)  Comments: |
| --- | --- |

NAME OF MODERATOR: __________________________ DATE REVIEWED:_______________________

H. **Questionnaire**

**Instructions:**

- The moderator will ask 5 out of the 13 following questions to each operator (individually) in the context of the AlereLAM proficiency run.
- For each correct item, the operator will obtain 1 point.

NAME OF OPERATOR: __________________________	DATE OF COMPLETION:_________________

| **AlereLAM Questions** | | **Answered correctly** | | **If NO add comment** |
| --- | --- | --- | --- | --- |
| 30. What is the intended use of AlereLAM? | | ❑YES | ❑NO |  |
| 31. What is the storage temperature of the test? | | ❑YES | ❑NO |  |
| 32. Can you describe all the components that are included in the AlereLAM pouch? | | ❑YES | ❑NO |  |
| 33. What kind of specimen is required for the AlereLAM test? | | ❑YES | ❑NO |  |
| 34. Which lighting condition should be avoided while using the test? | | ❑YES | ❑NO |  |
| 35. What would you do if the pouch of the device was not opened right before testing but more than 2 hours in advance i.e. contents exposed to ambient conditions? | | ❑YES | ❑NO |  |
| 36. What would you do if the expiry date of the test has passed? | | ❑YES | ❑NO |  |
| 37. What would you do if you realize the lot number on the pouch is not identical to the one on the test strips? | | ❑YES | ❑NO |  |
| 38. What would you do if the amount of urine that you transferred to the test is more than 60 ul? | | ❑YES | ❑NO |  |
| 39. What would you do if you dropped the urine container and it splashed on the AlereLAM test(s)? | | ❑YES | ❑NO |  |
| 40. What is the minimum time to wait before test result interpretation? | | ❑YES | ❑NO |  |
| 41. What is the maximum time within which you need to interpret the test results? | | ❑YES | ❑NO |  |
| 42. How should you dispose of the AlereLAM cartridge? | | ❑YES | ❑NO |  |
| **PART C** | **Score / Number of correct questions** | **/ 5** | | …………… % |

NAME OF MODERATOR: __________________________	DATE REVIEWED:_________________

I. **Test result interpretation**

**Instructions:**

- The moderator will provide a form containing 10 photographs of possible test results.
- The operator has to interpret the result for each case using the reference card of the AlereLAM test.
- For each correct item, the operator will obtain 1 point.

NAME OF OPERATOR: __________________________	DATE OF COMPLETION:_________________

|  | **Test result example** | **Result interpretation** | | | **Moderator’s comment** |
| --- | --- | --- | --- | --- | --- |
| 1 | 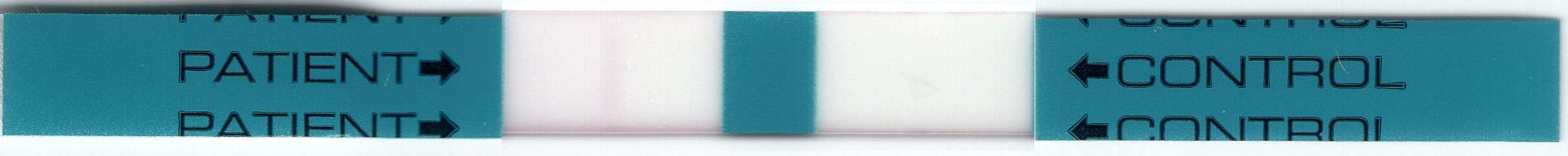 | ❑Positive  ❑1+  ❑2+  ❑3+  ❑4+ | ❑Negative | ❑Invalid |  |
| 2 | 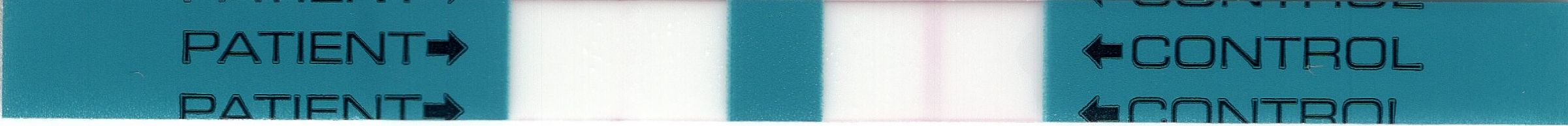 | ❑Positive  ❑1+  ❑2+  ❑3+  ❑4+ | ❑Negative | ❑Invalid |  |
| 3 | 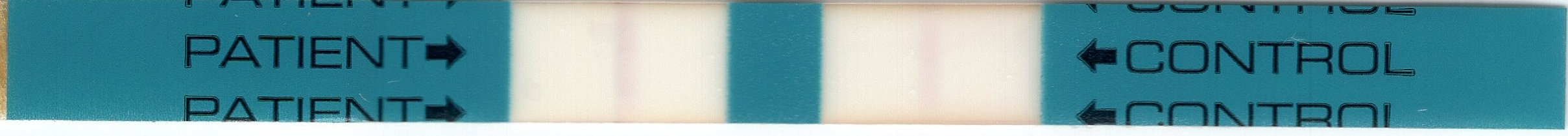 | ❑Positive  ❑1+  ❑2+  ❑3+  ❑4+ | ❑Negative | ❑Invalid |  |
| 4 | 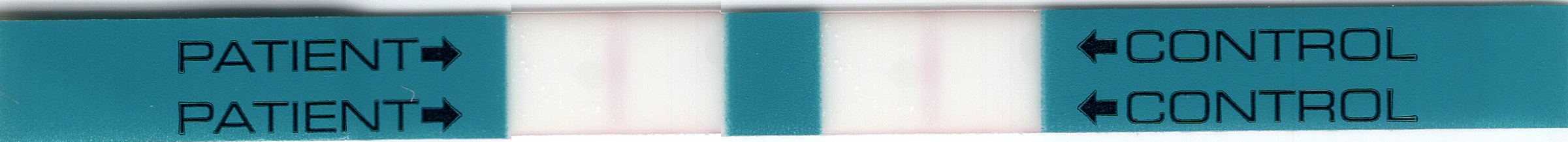 | ❑Positive  ❑1+  ❑2+  ❑3+  ❑4+ | ❑Negative | ❑Invalid |  |
| 5 | 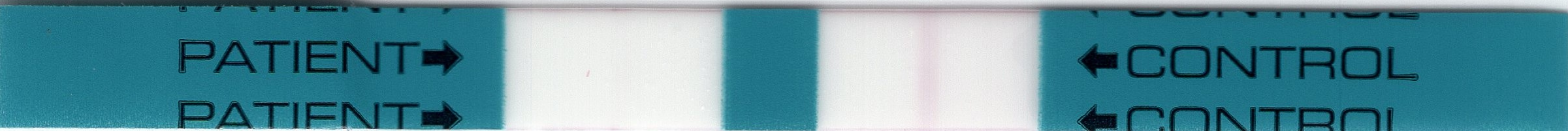 | ❑Positive  ❑1+  ❑2+  ❑3+  ❑4+ | ❑Negative | ❑Invalid |  |
| 6 | 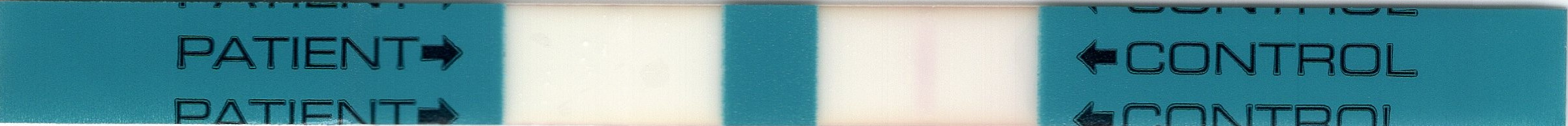 | ❑Positive  ❑1+  ❑2+  ❑3+  ❑4+ | ❑Negative | ❑Invalid |  |
| 7 | 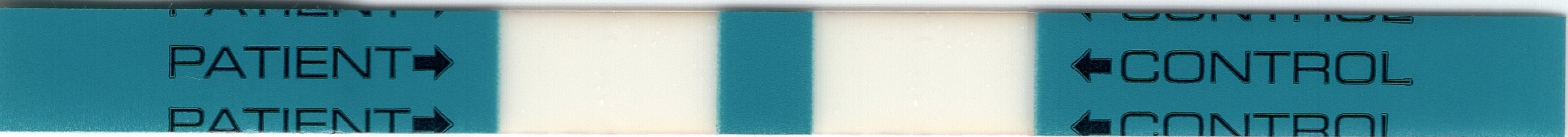 | ❑Positive  ❑1+  ❑2+  ❑3+  ❑4+ | ❑Negative | ❑Invalid |  |
| 8 | 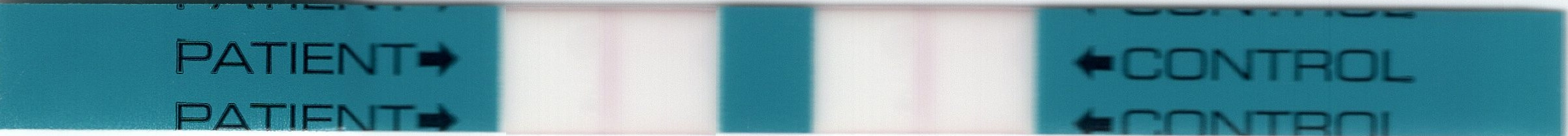 | ❑Positive  ❑1+  ❑2+  ❑3+  ❑4+ | ❑Negative | ❑Invalid |  |
| 9 | 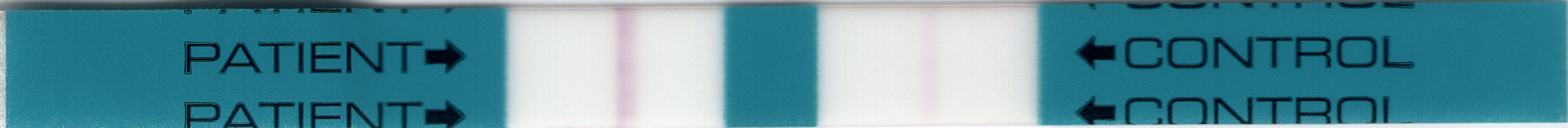 | ❑Positive  ❑1+  ❑2+  ❑3+  ❑4+ | ❑Negative | ❑Invalid |  |
| 10 | 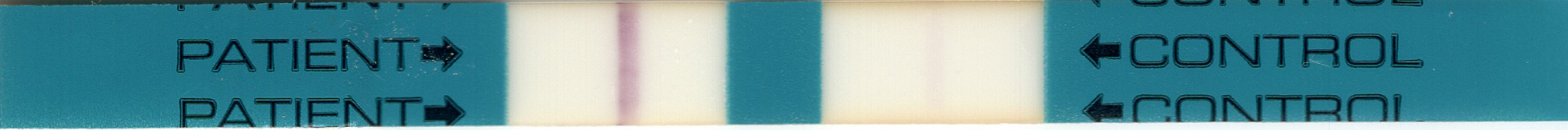 | ❑Positive  ❑1+  ❑2+  ❑3+  ❑4+ | ❑Negative | ❑Invalid |  |
| **PART D** | | **Score / Number of correct items** | | **/ 10** | …………… % |

J. **Conclusion**

| **Performance targets met?** |  |  | **If NO add comment** |
| --- | --- | --- | --- |
| Score Part A: ≥80%? (≥12) | ❑YES | ❑NO |  |
| Score Part B: Appraisal ≥4? | ❑YES | ❑NO |  |
| Score Part C: ≥80%? (≥10.4) | ❑YES | ❑NO |  |
| Score Part D: ≥90%? | ❑YES | ❑NO |  |
| **Conclusion: Operator passed proficiency test** | **❑YES**^#^ | **❑NO** |  |

^#^Operator can only pass the proficiency test, when score for individual Parts A, B, C and D were ALL met.

NAME OF MODERATOR: __________________________	DATE ASSESSED:_________________

**Figures and Tables**

**Figure E1. Study testing procedures**

A. **Day 1**

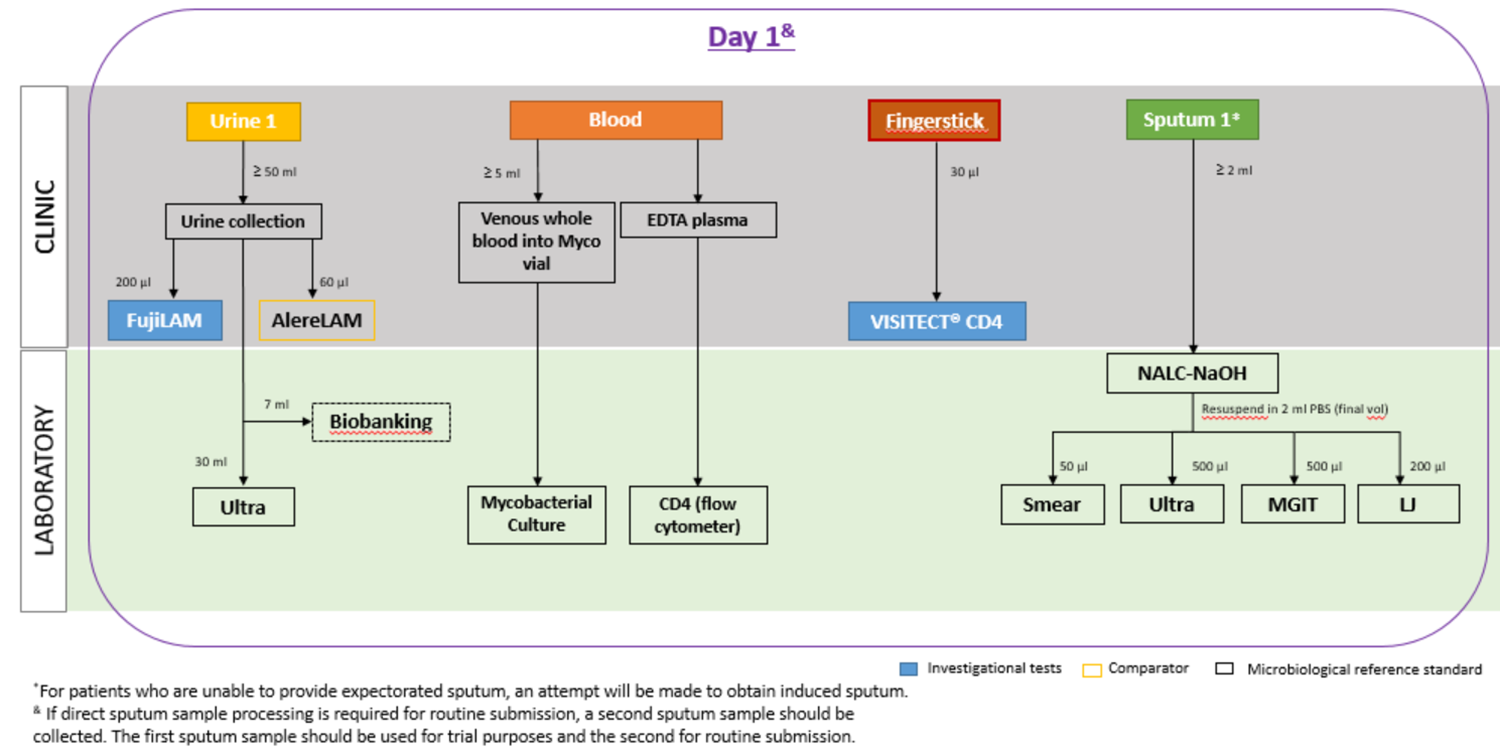

**B. Day 2**

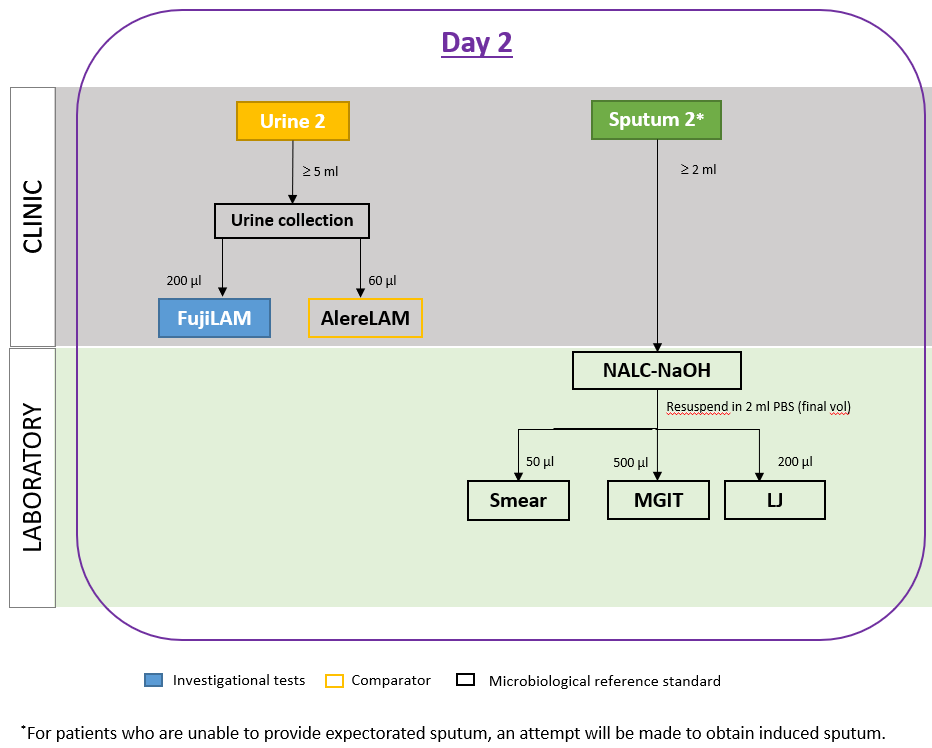

C.
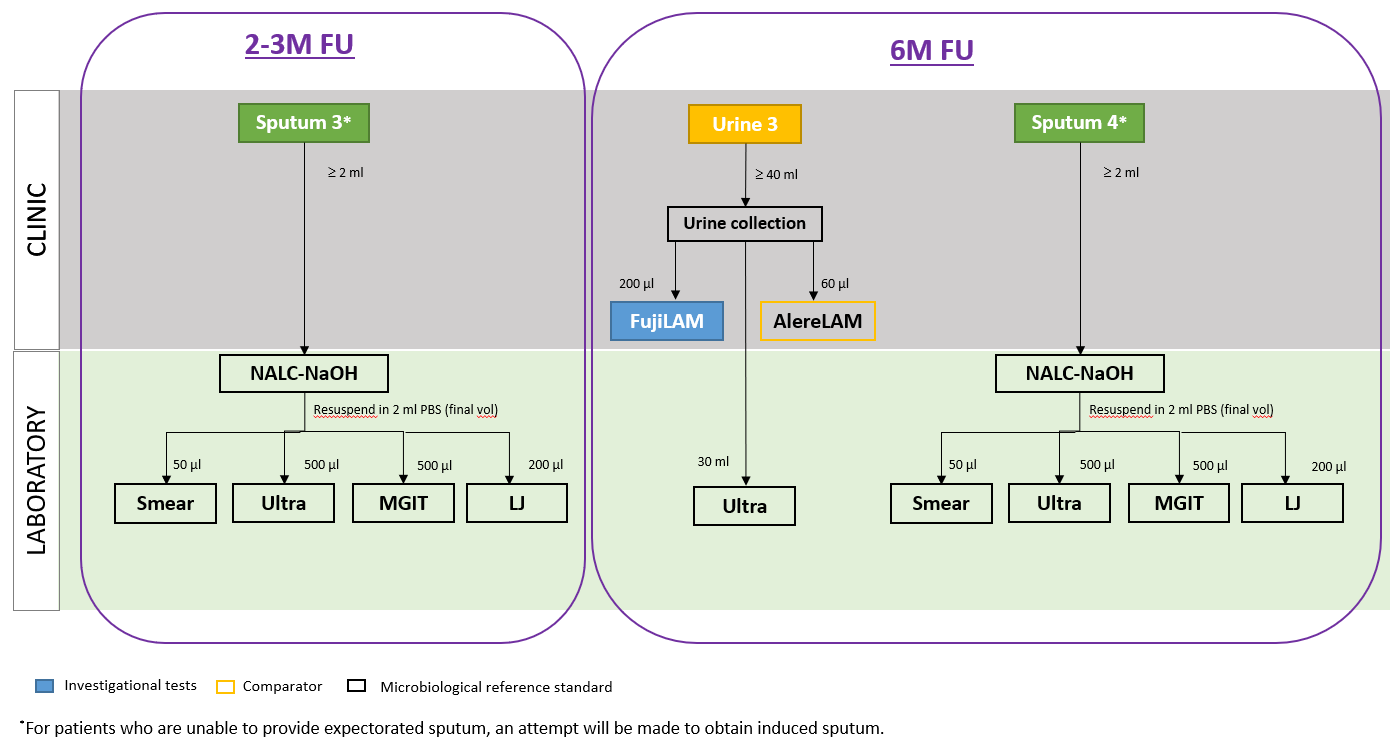
**Follow-up**

LJ, Löwenstein-Jensen; MGIT, Mycobacteria Growth Indicator Tube.

**Figure E2: Lot distribution across countries**

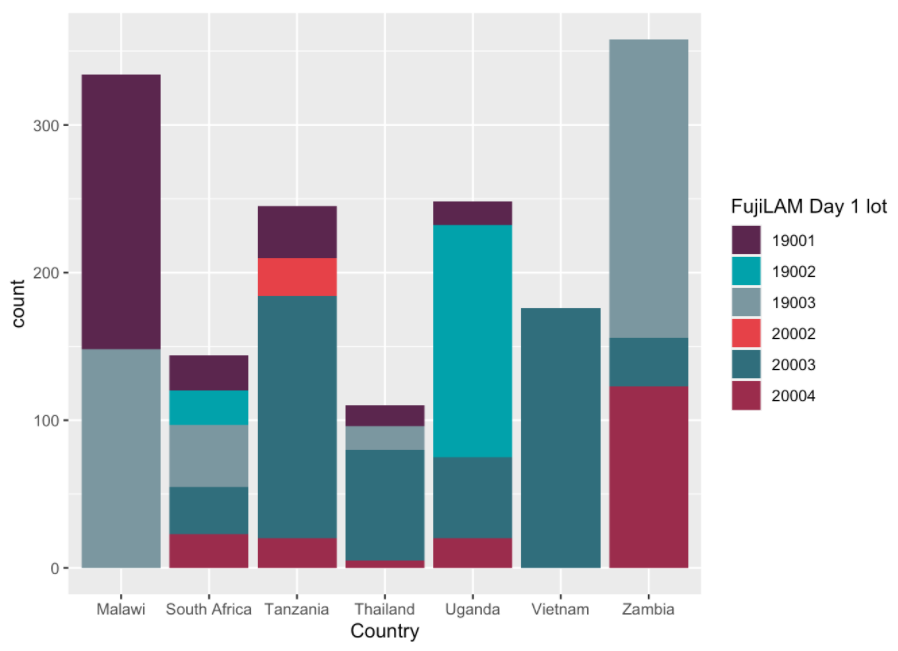

**Figure E3: Correlation of mismatch with CD4 cell count of TB positive cases**

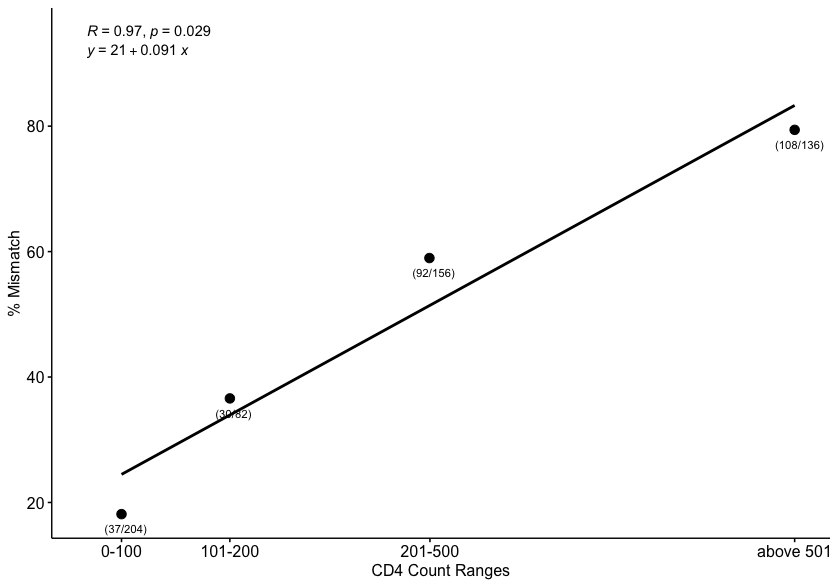

**Table E1. Participating centres, locations, settings, collection sites**

| **Country** | **Organization** | **Setting** | **Collection sites** | **Level of care** | **City** |
| --- | --- | --- | --- | --- | --- |
| **South Africa** | University of Cape Town | Inpatient | Mitchells Plain Hospital | District | Cape Town |
| **Malawi** | Malawi-Liverpool-Wellcome Programme and College of Medicine, University of Malawi | In- and outpatient | Queen Elizabeth Central Hospital | Regional referral | Blantyre |
|  |  |  | Bangwe Primary Care Clinic | Primary | Blantyre |
| **Zambia** | Centre for Infectious Disease Research in Zambia | In- and outpatient | Kanyama Hospital | Primary | Lusaka |
|  |  |  | Chawama Hospital | Primary | Lusaka |
| **Uganda** | Infectious Diseases Institute | In- and outpatient | Kisenyi Health center IV Kampala | Sub-district | Kampala |
|  |  |  | Mulago National Referral Hospital_Kiruddu | National Referral |  |
| **Tanzania** | Swiss TPH & Ifakara Health Institute | Outpatient | Temeke Regional Referral Hospital | Regional Referral | Dar es Salaam |
| **Viet Nam** | Viet Tiep Hospital/National Lung Hospital | In- and outpatient | Viet Tiep Hospital | Primary | Hai Phong |
| **Thailand** | The HIV Netherlands Australia Thailand Research | In- and outpatient | Chulalongkorn Hospital | Referral | Bangkok |
|  |  |  | Taksin Hospital | Referral | Bangkok |
|  |  |  | Bamrasnaradura Institute | Referral | Nonthaburi |
|  |  |  | Public Health Center 28 | Referral | Bangkok |

**Table E2. List of tests and reference standard definitions^⧫^**

|  |  | **MRS** | **eMRS** | **CRS*** |
| --- | --- | --- | --- | --- |
| 1–2 Sputum MGIT culture^Ω^ | | YES | YES | YES |
| 1–2 Sputum LJ culture^Ω^ | | YES | YES | YES |
| Blood culture^Ω^ | | YES | YES | YES |
| Urine Xpert Ultra | | YES | YES | YES |
| Sputum Xpert Ultra | | YES | YES | YES |
| Additional (non-study) testing^§^ | | NO | YES | YES |
| 2–3-month follow-up testing | | YES | YES | YES |
| Anti-TB therapy with response | | NO | NO | YES |

CRS, composite reference standard; eMRS, extended microbiological reference standard; LJ, Löwenstein-Jensen; MGIT, Mycobacteria Growth Indicator Tube; MRS, microbiological reference standard. MTB, *Mycobacterium tuberculosis;* NTM, nontuberculous mycobacteria.

^Ω^ Including MTB complex confirmation and NTM determination

^§^ Any additional mycobacterial culture and/or Xpert/Ultra from other samples (e.g., pleural fluid, tissue biopsy, etc.) performed based on routine clinical indication.

*Chest X-Ray, AlereLAM and smear results might be considered as part of the clinical decision-making (as per country routine).

⧫The respective reference standard is considered positive if any of those marked with “YES” are positive/apply. The MRS/eMRS is negative if none of the tests marked with “YES” are positive and at least one negative sputum culture is available. CRS is negative if none of those marked with “YES” are positive/apply and participant has no symptoms at 2–3-month follow-up. Unclassifiable is neither reference standard positive nor reference standard negative.

**Table E3. Number of urine samples per country selected for the post-hoc exploratory analysis and respective lots used in the prospective evaluation study**

| **Lot** | **19001** | **19002** | **19003** | **20002** | **20003** | **20004** | **Total (selected)** |
| --- | --- | --- | --- | --- | --- | --- | --- |
| **Malawi** | 16 | - | 1 | - | - | - | 17 |
| **Tanzania** | 15 | - | - | 2 | 3 | - | 20 |
| **Thailand** | 11 | - | 8 | - | - | - | 19 |
| **Zambia** | - | - | 38* | - | 4 | 14 | 55 |
| **Total** | 40 | 0 | 49 | 2 | 7 | 14 | 111 |

*One sample not tested on EclLAM due to insufficient volume.

**Table E4. Demographic and clinical characteristics of study participants stratified by country**

|  | **South Africa** | **Malawi** | **Zambia** | **Uganda** | **Tanzania** | **Viet Nam** | **Thailand** |
| --- | --- | --- | --- | --- | --- | --- | --- |
| N | 148 | 336 | 358 | 248 | 245 | 177 | 112 |
| Age median [min-max] (years) | 39 [21––64] | 40 [18––82] | 39 [18–70] | 39 [18–71] | 45 [18–75] | 43 [18–75] | 33 [18–68] |
| Female, no. (%) | 86/148 (58) | 188/336 (56) | 196/358 (55) | 146/248 (59) | 178/245 (73) | 37/177 (21) | 13/112 (12) |
| Median CD4 count - cells/µl [min-max] | 103.5 [0–1283] | 409 [7–1170] | 343 [3–1173] | 371 [2–3643] | 608.5 [18–1464] | 458 [10–1387] | 177 [0–1013] |
| Seriously ill*, no. (%) | 148/148 (100) | 7/336 (2) | 20/358 (6) | 28/248 (11) | 0/245 (0) | 13/177 (7) | 1/112 (1) |
| History of TB, no. (%) | 76/148 (51) | 62/336 (18) | 125/358 (35) | 39/248 (16) | 68/245 (28) | 52/177 (29) | 6/112 (5) |
| WHO TB Symptoms, no. (%) | 125/148 (84) | 312/336 (93) | 323/358 (90) | 241/248 (97) | 245/245 (100) | 156/177 (88) | 106/112 (95) |
| ***Setting*** |  |  |  |  |  |  |  |
| Inpatients, no. (%) | 148/148 (100) | 161/336 (48) | 141/358 (39) | 125/248 (50) | 0/245 (0) | 78/177 (44) | 25/112 (22) |
| Outpatients, no. (%) | 0/148 (0) | 175/336 (52) | 217/358 (61) | 123/248 (50) | 245/245 (100) | 99/177 (56) | 87/112 (78) |
| ***CD4*** |  |  |  |  |  |  |  |
| ≤100 | 71/148 (48) | 54/336 (16) | 53/358 (15) | 67/248 (27) | 15/245 (6) | 34/177 (19) | 36/112 (32) |
| >100 to ≤200 | 22/148 (15) | 43/336 (13) | 55/358 (15) | 17/248 (7) | 14/245 (6) | 9/177 (5) | 25/112 (22) |
| >200 to ≤500 | 28/148 (19) | 109/336 (32) | 143/358 (40) | 64/248 (26) | 60/245 (24) | 57/177 (32) | 42/112 (38) |
| >500 | 23/148 (16) | 129/336 (38) | 102/358 (28) | 99/248 (40) | 155/245 (63) | 75/177 (42) | 7/112 (6) |
| CD4 Unknown | 4/148 (3) | 1/336 (0) | 5/358 (1) | 1/248 (0) | 1/245 (0) | 2/177 (1) | 2/112 (2) |
| Seriously ill* - CD4 ≤100, no. (%) | 71/71 (100) | 1/54 (2) | 8/53 (15) | 11/67 (16) | 0/15 (0) | 11/34 (32) | 0/36 (0) |
| Seriously ill* - CD4 ≤200, no. (%) | 93/93 (100) | 3/97 (3) | 11/108 (10) | 13/84 (15) | 0/29 (0) | 11/43 (26) | 0/61 (0) |
| ***HIV treatment*** |  |  |  |  |  |  |  |
| ART interrupted, no. (%) | 42/148 (28) | 6/336 (2) | 2/358 (1) | 11/248 (4) | 5/245 (2) | 14/177 (8) | 8/112 (7) |
| Currently on ART, no. (%) | 68/148 (46) | 302/336 (90) | 315/358 (88) | 179/248 (72) | 212/245 (87) | 147/177 (83) | 40/112 (36) |
| Don’t know, no. (%) | 0/148 (0) | 0/336 (0) | 9/358 (3) | 3/248 (1) | 0/245 (0) | 0/177 (0) | 5/112 (4) |
| Never used, no. (%) | 38/148 (26) | 28/336 (8) | 32/358 (9) | 55/248 (22) | 28/245 (11) | 16/177 (9) | 59/112 (53) |
| ***Speciation*** |  |  |  |  |  |  |  |
| NTM, no. (%) | 1/148 (1) | 0/336 (0) | 14/358 (4) | 12/248 (5) | 67/245 (27) | 0/177 (0) | 9/112 (8) |
| NTM&MTBC, no. (%) | 0/148 (0) | 1/336 (0) | 2/358 (1) | 0/248 (0) | 3/245 (1) | 0/177 (0) | 0/112 (0) |
| MTBC, no. (%) | 19/148 (13) | 19/336 (6) | 40/358 (11) | 30/248 (12) | 16/245 (7) | 14/177 (8) | 6/112 (5) |
| Not done/contaminated | 128/148 (86) | 316/336 (94) | 302/358 (84) | 206/248 (83) | 159/245 (65) | 163/177 (92) | 97/112 (87) |
| ***Follow-up status*** |  |  |  |  |  |  |  |
| Died within 3 months, no. (%) | 21/148 (14) | 27/336 (8) | 32/358 (9) | 37/248 (15) | 0/245 (0) | 4/177 (2) | 5/112 (4) |
| Alive at 3 months FU, no. (%) | 70/148 (47) | 277/336 (82) | 256/358 (72) | 163/248 (66) | 245/245 (100) | 132/177 (75) | 95/112 (85) |
| Lost to follow-up, no. (%) | 17/148 (11) | 32/336 (10) | 33/358 (9) | 16/248 (6) | 0/245 (0) | 13/177 (7) | 12/112 (11) |
| No follow-up needed, no. (%) | 40/148 (27) | 0/336 (0) | 37/358 (10) | 32/248 (13) | 0/245 (0) | 28/177 (16) | 0/112 (0) |

*Seriously ill if any of the followings present: respiratory rate > 30 breaths/min, heart rate > 120 beats/min, body mass index [BMI] ≤ 18.5 kg/m2, systolic blood pressure < 90 mmHg or being unable to walk unaided

ART, antiretroviral therapy; FU, follow-up; MTBC, Mycobacterium tuberculosis complex; no. number; NTM, non-tuberculous mycobacteria; TB, tuberculosis; WHO, World Health Organization.

**Table E5. Sensitivity and specificity of Day 2 FujiLAM against the eMRS**

|  | **N** | **TP** | **FP** | **FN** | **TN** | **Sensitivity % [95%CI]** | **Specificity % [95%CI]** |
| --- | --- | --- | --- | --- | --- | --- | --- |
| All | 1604 | 149 | 241 | 141 | 1073 | 51·4 [45·6–57·1] | 81·7 [79·5–83·7] |
| ***CD4*** |  | | | | | | |
| ≤100 | 322 | 80 | 58 | 19 | 165 | 80·8 [72·0–87·4] | 74·0 [67·9–79·3] |
| 101 to ≤200 | 184 | 25 | 28 | 15 | 116 | 62·5 [47·0–75·8] | 80·6 [73·3–86·2] |
| 201 to ≤500 | 498 | 29 | 80 | 49 | 340 | 37·2 [27·3–48·3] | 81·0 [76·9–84·4] |
| >500 | 587 | 13 | 74 | 55 | 445 | 19·1 [11·5–30·0] | 85·7 [82·5–88·5] |
| Unknown | 13 | 2 | 1 | 3 | 7 | 40·0 [11·8–76·9] | 87·5 [52·9–97·8] |
| ***Setting*** |  | | | | | | |
| Inpatient | 665 | 92 | 85 | 50 | 438 | 64·8 [56·6–72·2] | 83·7 [80·3–86·7] |
| Outpatient | 939 | 57 | 156 | 91 | 635 | 38·5 [31·1–46·6] | 80·3 [77·4–82·9] |
| ***Country*** |  | | | | | | |
| South Africa | 142 | 41 | 31 | 14 | 56 | 74·5 [61·7–84·2] | 64·4 [53·9–73·6] |
| Malawi | 333 | 21 | 58 | 11 | 243 | 65·6 [48·3–79·6] | 80·7 [75·9–84·8] |
| Zambia | 351 | 29 | 62 | 23 | 237 | 55·8 [42·3–68·4] | 79·3 [74·3–83·5] |
| Uganda | 248 | 28 | 44 | 16 | 160 | 63·6 [48·9–76·2] | 78·4 [72·3–83·5] |
| Tanzania | 245 | 14 | 20 | 54 | 157 | 20·6 [12·7–31·6] | 88·7 [83·2–92·6] |
| Viet Nam | 175 | 11 | 2 | 22 | 140 | 33·3 [19·8–50·4] | 98·6 [95·0–99·6] |
| Thailand | 110 | 5 | 24 | 1 | 80 | 83·3 [43·6–97·0] | 76·9 [68·0–84·0] |

eMRS, extended microbiological reference standard; FN, false negative; FP, false positive; N, number; TN, true negative; true positive.

**Table E6. Sensitivity and specificity of Day 1 and Day 2 AlereLAM against eMRS**

| **Day 1** | **N** | **TP** | **FP** | **FN** | **TN** | **Sensitivity % [95%CI]** | **Specificity % [95%CI]** |
| --- | --- | --- | --- | --- | --- | --- | --- |
| All | 1615 | 89 | 123 | 203 | 1200 | 30·5 [25·5–36·0] | 90·7 [89·0–92·2] |
| ***CD4*** |  | | | | | |  |
| ≤100 | 327 | 55 | 36 | 46 | 190 | 54·5 [44·8–63·8] | 84·1 [78·7–88·3] |
| 101 to ≤200 | 183 | 14 | 15 | 26 | 128 | 35·0 [22·1–50·5] | 89·5 [83·4–93·5] |
| 201 to ≤500 | 500 | 13 | 32 | 65 | 390 | 16·7 [10·0–26·5] | 92·4 [89·5–94·6] |
| >500 | 589 | 6 | 39 | 62 | 482 | 8·8 [4·1–17·9] | 92·5 [89·9–94·5] |
| Unknown | 16 | 1 | 1 | 4 | 10 | 20·0 [3·6–62·5] | 90·9 [62·3–98·4] |
| ***Setting*** |  | | | | | |  |
| Inpatient | 672 | 60 | 65 | 84 | 463 | 41·7 [33·9–49·8] | 87·7 [84·6–90·2] |
| Outpatient | 943 | 29 | 58 | 119 | 737 | 19·6 [14·0–26·7] | 92·7 [90·7–94·3] |
| ***Country*** |  | | | | | |  |
| South Africa | 144 | 17 | 1 | 39 | 87 | 30·4 [19·9–43·3] | 98·9 [93·8–99·8] |
| Malawi | 335 | 14 | 22 | 18 | 281 | 43·8 [28·2–60·7] | 92·7 [89·2–95·2] |
| Zambia | 358 | 12 | 46 | 41 | 259 | 22·6 [13·5–35·5] | 84·9 [80·5–88·5] |
| Uganda | 248 | 23 | 28 | 21 | 176 | 52·3 [37·9–66·2] | 86·3 [80·9–90·3] |
| Tanzania | 244 | 13 | 8 | 55 | 168 | 19·1 [11·5–30·0] | 95·5 [91·3–97·7] |
| Vietnam | 177 | 7 | 10 | 26 | 134 | 21·2 [10·7–37·8] | 93·1 [87·7–96·2] |
| Thailand | 109 | 3 | 8 | 3 | 95 | 50·0 [18·8–81·2] | 92·2 [85·4–96·0] |
| **Day 2** | **N** | **TP** | **FP** | **FN** | **TN** | **Sensitivity % [95%CI]** | **Specificity % [95%CI]** |
| All | 1601 | 82 | 164 | 207 | 1148 | 28·4 [23·5–33·8] | 87·5 [85·6–89·2] |
| ***CD4*** |  | | | | | |  |
| ≤100 | 323 | 52 | 36 | 47 | 188 | 52·5 [42·8–62·1] | 83·9 [78·6–88·2] |
| 101 to ≤200 | 181 | 11 | 16 | 29 | 125 | 27·5 [16·1–42·8] | 88·7 [82·4–92·9] |
| 201 to ≤500 | 497 | 10 | 55 | 67 | 365 | 13 [7·2–22·3] | 86·9 [83·3–89·8] |
| >500 | 587 | 7 | 55 | 61 | 464 | 10·3 [5·1–19·8] | 89·4 [86·5–91·8] |
| Unknown | 13 | 2 | 2 | 3 | 6 | 40 [11·8–76·9] | 75 [40·9–92·8] |
| ***Setting*** |  | | | | | |  |
| Inpatient | 666 | 58 | 61 | 83 | 464 | 41·1 [33·4–49·4] | 88·4 [85·4–90·8] |
| Outpatient | 935 | 24 | 103 | 124 | 684 | 16·2 [11·2–23·0] | 86·9 [84·4–89·1] |
| ***Country*** |  | | | | | |  |
| South Africa | 142 | 19 | 1 | 36 | 86 | 34·5 [23·4–47·8] | 98·9 [93·8–99·8] |
| Malawi | 334 | 12 | 37 | 20 | 265 | 37·5 [22·9–54·8] | 87·7 [83·6–91·0] |
| Zambia | 351 | 14 | 58 | 38 | 241 | 26·9 [16·8–40·2] | 80·6 [75·7–84·7] |
| Uganda | 248 | 19 | 33 | 25 | 171 | 43·2 [29·7–57·8] | 83·8 [78·1–88·2] |
| Tanzania | 245 | 8 | 12 | 60 | 165 | 11·8 [6·1–21·5] | 93·2 [88·5–96·1] |
| Vietnam | 175 | 8 | 8 | 24 | 135 | 25 [13·2–42·1] | 94·4 [89·3–97·1] |
| Thailand | 106 | 2 | 15 | 4 | 85 | 33·3 [9·7–70·0] | 85 [76·7–90·7] |

eMRS, extended microbiological reference standard; FN, false negative; FP, false positive; N, number; TN, true negative; true positive.

**Table E7.** **Sensitivity and specificity of Day 1 and Day 2 FujiLAM against the CRS**

| **Day 1** | **N** | **TP** | **FP** | **FN** | **TN** | **Sensitivity % [95%CI]** | **Specificity % [95%CI]** |
| --- | --- | --- | --- | --- | --- | --- | --- |
| All | 1399 | 218 | 125 | 264 | 792 | 45·2 [40·8–49·7] | 86·4 [84·0–88·4] |
| ***CD4*** |  | | | | | | |
| ≤100 | 259 | 100 | 17 | 50 | 92 | 66·7 [58·8–73·7] | 84·4 [76·4–90·0] |
| 101 to ≤200 | 155 | 31 | 11 | 39 | 74 | 44·3 [33·2–55·9] | 87·1 [78·3–92·6] |
| 201 to ≤500 | 434 | 53 | 48 | 90 | 243 | 37·1 [29·6–45·2] | 83·5 [78·8–87·3] |
| >500 | 542 | 32 | 48 | 81 | 381 | 28·3 [20·8–37·2] | 88·8 [85·5–91·5] |
| Unknown | 9 | 2 | 1 | 4 | 2 | 33·3 [9·7–70·0] | 66·7 [20·8–93·8] |
| ***Setting*** |  | | | | | | |
| Inpatient | 520 | 126 | 26 | 111 | 257 | 53·2 [46·8–59·4] | 90·8 [86·9–93·7] |
| Outpatient | 879 | 92 | 99 | 153 | 535 | 37·6 [31·7–43·8] | 84·4 [81·3–87·0] |
| ***Country*** |  | | | | | | |
| South Africa | 137 | 46 | 16 | 23 | 52 | 66·7 [54·9–76·6] | 76·5 [65·1–85·0] |
| Malawi | 280 | 23 | 31 | 30 | 196 | 43·4 [31·0–56·7] | 86·3 [81·3–90·2] |
| Zambia | 303 | 59 | 25 | 76 | 143 | 43·7 [35·6–52·1] | 85·1 [79·0–89·7] |
| Uganda | 204 | 50 | 11 | 51 | 92 | 49·5 [40·0–59·1] | 89·3 [81·9–93·9] |
| Tanzania | 245 | 21 | 20 | 54 | 150 | 28·0 [19·1–39·0] | 88·2 [82·5–92·2] |
| Vietnam | 135 | 11 | 5 | 23 | 96 | 32·4 [19·1–49·2] | 95·0 [88·9–97·9] |
| Thailand | 95 | 8 | 17 | 7 | 63 | 53·3 [30·1–75·2] | 78·8 [68·6–86·3] |
| **Day 2** | **N** | **TP** | **FP** | **FN** | **TN** | **Sensitivity % [95%CI]** | **Specificity % [95%CI]** |
| All | 1393 | 210 | 153 | 269 | 761 | 43·8 [39·5–48·3] | 83·3 [80·7–85·5] |
| ***CD4*** |  | | | | | | |
| ≤100 | 257 | 101 | 26 | 47 | 83 | 68·2 [60·4–75·2] | 76·1 [67·3–83·2] |
| 101 to ≤200 | 155 | 32 | 15 | 38 | 70 | 45·7 [34·6–57·3] | 82·4 [72·9–89·0] |
| 201 to ≤500 | 431 | 46 | 55 | 96 | 234 | 32·4 [25·2–40·5] | 81 [76·0–85·1] |
| >500 | 541 | 29 | 56 | 84 | 372 | 25·7 [18·5–34·4] | 86·9 [83·4–89·8] |
| Unknown | 9 | 2 | 1 | 4 | 2 | 33·3 [9·7–70·0] | 66·7 [20·8–93·8] |
| ***Setting*** |  | | | | | | |
| Inpatient | 513 | 123 | 36 | 112 | 242 | 52·3 [46·0–58·6] | 87·1 [82·6–90·5] |
| Outpatient | 880 | 87 | 117 | 157 | 519 | 35·7 [29·9–41·9] | 81·6 [78·4–84·4] |
| ***Country*** |  | | | | | | |
| South Africa | 133 | 46 | 19 | 22 | 46 | 67·6 [55·8–77·6] | 70·8 [58·8–80·4] |
| Malawi | 279 | 28 | 42 | 25 | 184 | 52·8 [39·7–65·6] | 81·4 [75·8–86·0] |
| Zambia | 301 | 52 | 34 | 81 | 134 | 39·1 [31·2–47·6] | 79·8 [73·0–85·1] |
| Uganda | 204 | 49 | 16 | 52 | 87 | 48·5 [39·0–58·1] | 84·5 [76·2–90·2] |
| Tanzania | 245 | 16 | 18 | 59 | 152 | 21·3 [13·6–31·9] | 89·4 [83·9–93·2] |
| Vietnam | 135 | 11 | 2 | 23 | 99 | 32·4 [19·1–49·2] | 98 [93·1–99·5] |
| Thailand | 96 | 8 | 22 | 7 | 59 | 53·3 [30·1–75·2] | 72·8 [62·3–81·3] |

CRS, composite reference standard; FN, false negative; FP, false positive; N, number; TN, true negative; true positive.

**Table E8. Sensitivity and specificity of Day 1 and Day 2 AlereLAM against CRS**

| **Day 1** | **N** | **TP** | **FP** | **FN** | **TN** | **Sensitivity % [95%CI]** | **Specificity % [95%CI]** |
| --- | --- | --- | --- | --- | --- | --- | --- |
| All | 1398 | 146 | 43 | 336 | 873 | 30·3 [26·4–34·5] | 95·3 [93·7–96·5] |
| ***CD4*** |  | | | | | | |
| ≤100 | 259 | 74 | 7 | 76 | 102 | 49·3 [41·4–57·2] | 93·6 [87·3–96·9] |
| 101 to ≤200 | 155 | 20 | 7 | 50 | 78 | 28·6 [19·3–40·1] | 91·8 [84·0–96·0] |
| 201 to ≤500 | 432 | 29 | 8 | 114 | 281 | 20·3 [14·5–27·6] | 97·2 [94·6–98·6] |
| >500 | 543 | 22 | 21 | 91 | 409 | 19·5 [13·2–27·7] | 95·1 [92·7–96·8] |
| Unknown | 9 | 1 | 0 | 5 | 3 | 16·7 [3·0–56·4] | 100·0 [43·9–100·0] |
| ***Setting*** |  | | | | | | |
| Inpatient | 520 | 92 | 13 | 145 | 270 | 38·8 [32·8–45·1] | 95·4 [92·3–97·3] |
| Outpatient | 878 | 54 | 30 | 191 | 603 | 22·0 [17·3–27·6] | 95·3 [93·3–96·7] |
| ***Country*** |  | | | | | | |
| South Africa | 137 | 21 | 1 | 48 | 67 | 30·4 [20·8–42·1] | 98·5 [92·1–99·7] |
| Malawi | 280 | 18 | 16 | 35 | 211 | 34·0 [22·7–47·4] | 93·0 [88·9–95·6] |
| Zambia | 303 | 41 | 9 | 94 | 159 | 30·4 [23·2–38·6] | 94·6 [90·1–97·2] |
| Uganda | 204 | 40 | 3 | 61 | 100 | 39·6 [30·6–49·4] | 97·1 [91·8–99·0] |
| Tanzania | 244 | 14 | 7 | 61 | 162 | 18·7 [11·5–28·9] | 95·9 [91·7–98·0] |
| Vietnam | 136 | 7 | 5 | 27 | 97 | 20·6 [10·3–36·8] | 95·1 [89·0–97·9] |
| Thailand | 94 | 5 | 2 | 10 | 77 | 33·3 [15·2–58·3] | 97·5 [91·2–99·3] |
| **Day 2** | **N** | **TP** | **FP** | **FN** | **TN** | **Sensitivity % [95%CI]** | **Specificity % [95%CI]** |
| All | 1388 | 138 | 80 | 341 | 829 | 28·8 [24·9–33·0] | 91·2 [89·2–92·9] |
| ***CD4*** |  | | | | | | |
| ≤100 | 257 | 67 | 8 | 81 | 101 | 45·3 [37·5–53·3] | 92·7 [86·2–96·2] |
| 101 to ≤200 | 152 | 17 | 6 | 53 | 76 | 24·3 [15·8–35·5] | 92·7 [84·9–96·6] |
| 201 to ≤500 | 429 | 32 | 28 | 110 | 259 | 22·5 [16·4–30·1] | 90·2 [86·3–93·2] |
| >500 | 541 | 20 | 37 | 93 | 391 | 17·7 [11·8–25·8] | 91·4 [88·3–93·7] |
| Unknown | 9 | 2 | 1 | 4 | 2 | 33·3 [9·7–70·0] | 66·7 [20·8–93·8] |
| ***Setting*** |  | | | | | | |
| Inpatient | 513 | 87 | 11 | 147 | 268 | 37·2 [31·2–43·5] | 96·1 [93·1–97·8] |
| Outpatient | 875 | 51 | 69 | 194 | 561 | 20·8 [16·2–26·3] | 89 [86·4–91·2] |
| ***Country*** |  | | | | | | |
| South Africa | 133 | 21 | 0 | 47 | 65 | 30·9 [21·2–42·6] | 100 [94·4–100·0] |
| Malawi | 279 | 16 | 31 | 37 | 195 | 30·2 [19·5–43·5] | 86·3 [81·2–90·2] |
| Zambia | 301 | 46 | 16 | 88 | 151 | 34·3 [26·8–42·7] | 90·4 [85·0–94·0] |
| Uganda | 204 | 34 | 8 | 67 | 95 | 33·7 [25·2–43·3] | 92·2 [85·4–96·0] |
| Tanzania | 245 | 8 | 12 | 67 | 158 | 10·7 [5·5–19·7] | 92·9 [88·1–95·9] |
| Vietnam | 135 | 8 | 6 | 25 | 96 | 24·2 [12·8–41·0] | 94·1 [87·8–97·3] |
| Thailand | 91 | 5 | 7 | 10 | 69 | 33·3 [15·2–58·3] | 90·8 [82·2–95·5] |

CRS, composite reference standard; FN, false negative; FP, false positive; N, number; TN, true negative; true positive.

**Table E9.** **Sensitivity and specificity of Day 1 and Day 2 FujiLAM against the MRS**

| **Day 1** | **N** | **TP** | **FP** | **FN** | **TN** | **Sensitivity % [95%CI]** | **Specificity % [95%CI]** |
| --- | --- | --- | --- | --- | --- | --- | --- |
| All | 1609 | 156 | 197 | 129 | 1127 | 54·7 [48·9–60·4] | 85·1 [83·1–86·9] |
| ***CD4*** |  | | | | | | |
| ≤100 | 321 | 79 | 44 | 17 | 181 | 82·3 [73·5–88·6] | 80·4 [74·8–85·1] |
| 101 to ≤200 | 183 | 25 | 18 | 15 | 125 | 62·5 [47·0–75·8] | 87·4 [81·0–91·9] |
| 201 to ≤500 | 502 | 35 | 70 | 43 | 354 | 44·9 [34·3–55·9] | 83·5 [79·7–86·7] |
| >500 | 588 | 15 | 64 | 52 | 457 | 22·4 [14·1–33·7] | 87·7 [84·6–90·3] |
| Unknown | 15 | 2 | 1 | 2 | 10 | 50 [15·0–85·0] | 90·9 [62·3–98·4] |
| ***Setting*** |  | | | | | | |
| Inpatient | 665 | 96 | 63 | 41 | 465 | 70·1 [61·9–77·1] | 88·1 [85·0–90·6] |
| Outpatient | 944 | 60 | 134 | 88 | 662 | 40·5 [33·0–48·6] | 83·2 [80·4–85·6] |
| ***Country*** |  | | | | | | |
| South Africa | 138 | 37 | 22 | 12 | 67 | 75·5 [61·9–85·4] | 75·3 [65·4–83·1] |
| Malawi | 334 | 20 | 36 | 12 | 266 | 62·5 [45·2–77·1] | 88·1 [83·9–91·3] |
| Zambia | 358 | 33 | 56 | 20 | 249 | 62·3 [48·8–74·1] | 81·6 [76·9–85·6] |
| Uganda | 248 | 32 | 36 | 12 | 168 | 72·7 [58·1–83·7] | 82·4 [76·5–87·0] |
| Tanzania | 245 | 18 | 23 | 50 | 154 | 26·5 [17·4–38·0] | 87·0 [81·3–91·2] |
| Vietnam | 176 | 11 | 5 | 22 | 138 | 33·3 [19·8–50·4] | 96·5 [92·1–98·5] |
| Thailand | 110 | 5 | 19 | 1 | 85 | 83·3 [43·6–97·0] | 81·7 [73·2–88·0] |
| **Day 2** | **N** | **TP** | **FP** | **FN** | **TN** | **Sensitivity % [95%CI]** | **Specificity % [95%CI]** |
| All | 1598 | 145 | 241 | 138 | 1074 | 51·2 [45·4–57·0] | 81·7 [79·5–83·7] |
| ***CD4*** |  | | | | | | |
| ≤100 | 317 | 76 | 58 | 18 | 165 | 80·9 [71·8–87·5] | 74 [67·9–79·3] |
| 101 to ≤200 | 184 | 25 | 28 | 15 | 116 | 62·5 [47·0–75·8] | 80·6 [73·3–86·2] |
| 201 to ≤500 | 498 | 29 | 80 | 49 | 340 | 37·2 [27·3–48·3] | 81 [76·9–84·4] |
| >500 | 587 | 13 | 74 | 54 | 446 | 19·4 [11·7–30·4] | 85·8 [82·5–88·5] |
| Unknown | 12 | 2 | 1 | 2 | 7 | 50 [15·0–85·0] | 87·5 [52·9–97·8] |
| ***Setting*** |  | | | | | | |
| Inpatient | 659 | 88 | 85 | 47 | 439 | 65·2 [56·8–72·7] | 83·8 [80·4–86·7] |
| Outpatient | 939 | 57 | 156 | 91 | 635 | 38·5 [31·1–46·6] | 80·3 [77·4–82·9] |
| ***Country*** |  | | | | | | |
| South Africa | 136 | 37 | 31 | 11 | 57 | 77·1 [63·5–86·7] | 64·8 [54·4–73·9] |
| Malawi | 333 | 21 | 58 | 11 | 243 | 65·6 [48·3–79·6] | 80·7 [75·9–84·8] |
| Zambia | 351 | 29 | 62 | 23 | 237 | 55·8 [42·3–68·4] | 79·3 [74·3–83·5] |
| Uganda | 248 | 28 | 44 | 16 | 160 | 63·6 [48·9–76·2] | 78·4 [72·3–83·5] |
| Tanzania | 245 | 14 | 20 | 54 | 157 | 20·6 [12·7–31·6] | 88·7 [83·2–92·6] |
| Vietnam | 175 | 11 | 2 | 22 | 140 | 33·3 [19·8–50·4] | 98·6 [95·0–99·6] |
| Thailand | 106 | 2 | 15 | 4 | 85 | 33·3 [9·7–70·0] | 85 [76·7–90·7] |

MRS, microbiological reference standard; FN, false negative; FP, false positive; N, number; TN, true negative; true positive.

**Table E10.** **Sensitivity and specificity of Day 1 and Day 2 AlereLAM against the MRS**

| **Day 1** | **N** | **TP** | **FP** | **FN** | **TN** | **Sensitivity [95%CI]** | **Specificity [95%CI]** |
| --- | --- | --- | --- | --- | --- | --- | --- |
| All | 1609 | 87 | 123 | 198 | 1201 | 30·5 [25·5–36·1] | 90·7 [89·0–92·2] |
| ***CD4*** |  | | | | | |  |
| ≤100 | 322 | 53 | 36 | 43 | 190 | 55·2 [45·2–64·8] | 84·1 [78·7–88·3] |
| 101 to ≤200 | 183 | 14 | 15 | 26 | 128 | 35·0 [22·1–50·5] | 89·5 [83·4–93·5] |
| 201 to ≤500 | 500 | 13 | 32 | 65 | 390 | 16·7 [10·0–26·5] | 92·4 [89·5–94·6] |
| >500 | 589 | 6 | 39 | 61 | 483 | 9 [4·2–18·2] | 92·5 [90·0–94·5] |
| Unknown | 15 | 1 | 1 | 3 | 10 | 25 [4·6–69·9] | 90·9 [62·3–98·4] |
| ***Setting*** |  | | | | | |  |
| Inpatient | 666 | 58 | 65 | 79 | 464 | 42·3 [34·4–50·7] | 87·7 [84·6–90·2] |
| Outpatient | 943 | 29 | 58 | 119 | 737 | 19·6 [14·0–26·7] | 92·7 [90·7–94·3] |
| ***Country*** |  | | | | | |  |
| South Africa | 138 | 15 | 1 | 34 | 88 | 30·6 [19·5–44·5] | 98·9 [93·9–99·8] |
| Malawi | 335 | 14 | 22 | 18 | 281 | 43·8 [28·2–60·7] | 92·7 [89·2–95·2] |
| Zambia | 358 | 12 | 46 | 41 | 259 | 22·6 [13·5–35·5] | 84·9 [80·5–88·5] |
| Uganda | 248 | 23 | 28 | 21 | 176 | 52·3 [37·9–66·2] | 86·3 [80·9–90·3] |
| Tanzania | 244 | 13 | 8 | 55 | 168 | 19·1 [11·5–30·0] | 95·5 [91·3–97·7] |
| Vietnam | 177 | 7 | 10 | 26 | 134 | 21·2 [10·7–37·8] | 93·1 [87·7–96·2] |
| Thailand | 109 | 3 | 8 | 3 | 95 | 50·0 [18·8–81·2] | 92·2 [85·4–96·0] |
| **Day 2** | **N** | **TP** | **FP** | **FN** | **TN** | **Sensitivity [95%CI]** | **Specificity [95%CI]** |
| All | 1595 | 80 | 164 | 202 | 1149 | 28·4 [23·4–33·9] | 87·5 [85·6–89·2] |
| ***CD4*** |  | | | | | |  |
| ≤100 | 318 | 50 | 36 | 44 | 188 | 53·2 [43·2–62·9] | 83·9 [78·6–88·2] |
| 101 to ≤200 | 181 | 11 | 16 | 29 | 125 | 27·5 [16·1–42·8] | 88·7 [82·4–92·9] |
| 201 to ≤500 | 497 | 10 | 55 | 67 | 365 | 13 [7·2–22·3] | 86·9 [83·3–89·8] |
| >500 | 587 | 7 | 55 | 60 | 465 | 10·4 [5·1–20·0] | 89·4 [86·5–91·8] |
| Unknown | 12 | 2 | 2 | 2 | 6 | 50 [15·0–85·0] | 75 [40·9–92·8] |
| ***Setting*** |  | | | | | |  |
| Inpatient | 660 | 56 | 61 | 78 | 465 | 41·8 [33·8–50·3] | 88·4 [85·4–90·9] |
| Outpatient | 935 | 24 | 103 | 124 | 684 | 16·2 [11·2–23·0] | 86·9 [84·4–89·1] |
| ***Country*** |  | | | | | |  |
| South Africa | 136 | 17 | 1 | 31 | 87 | 35·4 [23·4–49·6] | 98·9 [93·8–99·8] |
| Malawi | 334 | 12 | 37 | 20 | 265 | 37·5 [22·9–54·8] | 87·7 [83·6–91·0] |
| Zambia | 351 | 14 | 58 | 38 | 241 | 26·9 [16·8–40·2] | 80·6 [75·7–84·7] |
| Uganda | 248 | 19 | 33 | 25 | 171 | 43·2 [29·7–57·8] | 83·8 [78·1–88·2] |
| Tanzania | 245 | 8 | 12 | 60 | 165 | 11·8 [6·1–21·5] | 93·2 [88·5–96·1] |
| Vietnam | 175 | 8 | 8 | 24 | 135 | 25 [13·2–42·1] | 94·4 [89·3–97·1] |
| Thailand | 106 | 2 | 15 | 4 | 85 | 33·3 [9·7–70·0] | 85 [76·7–90·7] |

MRS, microbiological reference standard; FN, false negative; FP, false positive; N, number; TN, true negative; true positive.

**Table E11. Additional microbiological non-study tests contributing to eMRS**

|  | **South Africa** | **Malawi** | **Zambia** | **Uganda** | **Tanzania** | **Viet Nam** | **Thailand** |
| --- | --- | --- | --- | --- | --- | --- | --- |
| **Additional mycobacterial culture and/or Xpert Ultra from additional samples performed based on routine clinical indication and considered for the eMRS classification** | 123/199 | 0/349 | 13/360 | 0/249 | 0/245 | 3/177 | 1/113 |

eMRS, extended microbiological reference standard.

**Table E12. Summary statistics for the generalized linear mixed model fit on the mismatch ratio between FujiLAM and the reference of the full dataset**

| **Country** | **Chisq** | **Df** | **Pr (>Chisq)** |
| --- | --- | --- | --- |
| ***Age*** | 0·201 | 1 | 0·654 |
| ***Sex*** | 0·91 | 1 | 0·34 |
| ***Country*** | 38·807 | 6 | 7·81e-07 |
| ***Lot*** | 51·4 | 5 | 7·16e-10 |
| ***Visit day*** | 9·095 | 1 | 0·003 |
| ***logCD4*** | 0·101 | 1 | 0·75 |
| ***Urine color*** | 0·882 | 4 | 0·927 |
| ***Urine turbidity*** | 1·73 | 4 | 0·785 |
| ***Setting*** | 2·655 | 1 | 0·103 |

Chisq, chi-squared test; Df, degrees of freedom; Pr, probability.

**Table E13. Detailed results of the generalized linear mix model fit on the mismatch ratio between FujiLAM and the reference on the full dataset**

|  |  | **Estimate** | **Std. Error** | **z value** | **Pr(>\|z\|)** | **OR** | **2·5 %** | **97·5 %** |
| --- | --- | --- | --- | --- | --- | --- | --- | --- |
| (Intercept) |  | 0·031 | 0·822 | 0·037 | 0·97 | 1·031 | 0·206 | 5·163 |
| ***Age*** |  | 0·005 | 0·01 | 0·449 | 0·654 | 1·005 | 0·984 | 1·025 |
| ***Sex*** | *Male* | 0·229 | 0·24 | 0·954 | 0·34 | 1·258 | 0·785 | 2·015 |
| ***Country*** | *Malawi* | 2·05 | 0·491 | 4·174 | 2·99e-05 | 7·765 | 2·966 | 20·33 |
|  | *Zambia* | 0·819 | 0·492 | 1·665 | 0·096 | 2·267 | 0·865 | 5·945 |
|  | *Uganda* | 0·497 | 0·58 | 0·857 | 0·391 | 1·644 | 0·527 | 5·129 |
|  | *Tanzania* | -0·786 | 0·592 | -1·327 | 0·184 | 0·455 | 0·143 | 1·455 |
|  | *Vietnam* | -0·284 | 0·628 | -0·453 | 0·651 | 0·753 | 0·22 | 2·574 |
|  | *Thailand* | 0·329 | 0·656 | 0·501 | 0·616 | 1·389 | 0·384 | 5·023 |
| ***Lot number*** | *19002* | 1·126 | 0·568 | 1·983 | 0·047 | 3·084 | 1·013 | 9·389 |
|  | *19003* | 0·452 | 0·377 | 1·2 | 0·23 | 1·571 | 0·751 | 3·289 |
|  | *20002* | 3·765 | 1·003 | 3·755 | 1·73e-04 | 43·168 | 6·049 | 308·071 |
|  | *20003* | 2·793 | 0·431 | 6·481 | 9·13e-11 | 16·324 | 7·015 | 37·987 |
|  | *20004* | 1·75 | 0·492 | 3·557 | 3·74e-04 | 5·752 | 2·194 | 15·081 |
| ***Visit day*** | *Day 2* | -0·411 | 0·136 | -3·016 | 0·003 | 0·663 | 0·507 | 0·866 |
| ***CD4*** | *logCD4* | 0·073 | 0·229 | 0·319 | 0·75 | 1·076 | 0·686 | 1·686 |
| ***Urine Color*** | *Clear* | -0·012 | 0·449 | -0·027 | 0·978 | 0·988 | 0·41 | 2·381 |
|  | *Dark yellow* | 0·077 | 0·43 | 0·18 | 0·857 | 1·08 | 0·465 | 2·507 |
|  | *Light yellow* | -0·118 | 0·418 | -0·283 | 0·777 | 0·889 | 0·392 | 2·014 |
|  | *Red* | 16·712 | 1036·024 | 0·016 | 0·987 | 1·81e+07 | 0 |  |
| ***Urine Turbidity*** | *Cloudy* | -0·124 | 0·338 | -0·367 | 0·714 | 0·883 | 0·455 | 1·713 |
|  | *Flocculent* | 16·705 | 1348·11 | 0·012 | 0·99 | 1·80e+07 | 0 |  |
|  | *Opaque* | 0·093 | 0·806 | 0·115 | 0·908 | 1·097 | 0·226 | 5·326 |
|  | *Slightly cloudy* | 0·235 | 0·213 | 1·104 | 0·27 | 1·265 | 0·833 | 1·921 |
| ***Setting*** | *Outpatient* | -0·489 | 0·3 | -1·629 | 0·103 | 0·613 | 0·341 | 1·104 |

Std., standard; OR, odds ratio.

**Table E14. Summary statistics for the generalized linear mixed model fit on the mismatch ratio between FujiLAM and the reference on eMRS positives**

|  | **Chisq** | **Df** | **Pr (>Chisq)** |
| --- | --- | --- | --- |
| ***Age*** | 0·132 | 1 | 0·716 |
| ***Sex*** | 0·02 | 1 | 0·887 |
| ***Country*** | 4·143 | 6 | 0·657 |
| ***Lot*** | 5·696 | 5 | 0·337 |
| ***Visit day*** | 1·156 | 1 | 0·282 |
| ***logCD4*** | 20·73 | 1 | 5·29e-06 |
| ***Urine color*** | 2·474 | 4 | 0·649 |
| ***Urine turbidity*** | 1·261 | 3 | 0·738 |
| ***Setting*** | 0·066 | 1 | 0·797 |

Chisq, chi-squared test; Df, degrees of freedom; Pr, probability.

**Table E15. Detailed results of the generalized linear mix model fit on the mismatch ratio between FujiLAM and the reference on eMRS positives**

|  |  | **Estimate** | **Std. Error** | **z value** | **Pr(>\|z\|)** | **OR** | **2·5 %** | **97·5 %** |
| --- | --- | --- | --- | --- | --- | --- | --- | --- |
| (Intercept) |  | 9·326 | 2·235 | 4·172 | 3·02e-05 | 1·12e+04 | 140·483 | 8·97e+05 |
| ***Age*** |  | -0·011 | 0·032 | -0·364 | 0·716 | 0·989 | 0·929 | 1·052 |
| ***Sex*** | *Male* | 0·09 | 0·635 | 0·142 | 0·887 | 1·095 | 0·316 | 3·796 |
| ***Country*** | *Malawi* | -0·461 | 1·241 | -0·371 | 0·71 | 0·631 | 0·055 | 7·18 |
|  | *Zambia* | 0·179 | 1·098 | 0·163 | 0·87 | 1·196 | 0·139 | 10·279 |
|  | *Uganda* | 0·435 | 1·379 | 0·315 | 0·752 | 1·545 | 0·103 | 23·072 |
|  | *Tanzania* | -1·425 | 1·329 | -1·072 | 0·283 | 0·24 | 0·018 | 3·252 |
|  | *Vietnam* | -0·929 | 1·324 | -0·702 | 0·483 | 0·395 | 0·029 | 5·287 |
|  | *Thailand* | 2·042 | 2·344 | 0·871 | 0·384 | 7·71 | 0·078 | 762·232 |
| ***Lot number*** | *19002* | -0·801 | 1·472 | -0·544 | 0·586 | 0·449 | 0·025 | 8·044 |
|  | *19003* | -1·995 | 1·084 | -1·84 | 0·066 | 0·136 | 0·016 | 1·138 |
|  | *20002* | -0·211 | 2·626 | -0·08 | 0·936 | 0·81 | 0·005 | 139·12 |
|  | *20003* | -2·041 | 0·99 | -2·063 | 0·039 | 0·13 | 0·019 | 0·903 |
|  | *20004* | -2·104 | 1·203 | -1·749 | 0·08 | 0·122 | 0·012 | 1·29 |
| ***Visit day*** | *Day 2* | -0·365 | 0·339 | -1·075 | 0·282 | 0·694 | 0·357 | 1·35 |
| ***CD4*** | *logCD4* | -3·097 | 0·68 | -4·553 | 5·29e-06 | 0·045 | 0·012 | 0·171 |
| ***Urine Color*** | *Clear* | -0·509 | 1·01 | -0·504 | 0·614 | 0·601 | 0·083 | 4·355 |
|  | *Dark yellow* | 0·475 | 0·894 | 0·531 | 0·595 | 1·608 | 0·279 | 9·284 |
|  | *Light yellow* | -0·245 | 0·852 | -0·288 | 0·774 | 0·783 | 0·147 | 4·155 |
|  | *Red* | 18·855 | 2671·265 | 0·007 | 0·994 | 1·54e+08 | 0 |  |
| ***Urine Turbidity*** | *Cloudy* | 0·05 | 0·809 | 0·061 | 0·951 | 1·051 | 0·215 | 5·133 |
|  | *Opaque* | 0·044 | 1·447 | 0·03 | 0·976 | 1·045 | 0·061 | 17·816 |
|  | *Slightly cloudy* | 0·583 | 0·556 | 1·048 | 0·295 | 1·791 | 0·602 | 5·329 |
| ***Setting*** | *Outpatient* | 0·211 | 0·82 | 0·257 | 0·797 | 1·234 | 0·248 | 6·154 |

Std., standard; OR, odds ratio.

**Table E16. Summary statistics for the generalized linear mixed model fit on the mismatch ratio between FujiLAM and the reference of eMRS negatives**

|  | **Chisq** | **Df** | **Pr (>Chisq)** |
| --- | --- | --- | --- |
| ***Age*** | 0·063 | 1 | 0·801 |
| ***Sex*** | 6·371 | 1 | 0·012 |
| ***Country*** | 35·864 | 6 | 2·93e-06 |
| ***Lot*** | 97·232 | 5 | 2·02e-19 |
| ***Visit day*** | 7·094 | 1 | 0·008 |
| ***logCD4*** | 8·133 | 1 | 0·004 |
| ***Urine color*** | 0·176 | 4 | 0·996 |
| ***Urine turbidity*** | 0·986 | 4 | 0·912 |
| ***Setting*** | 7·792 | 1 | 0·005 |

Chisq, chi-squared test; Df, degrees of freedom; Pr, probability.

**Table E17. Detailed results of the generalized linear mix model fit on the mismatch ratio between FujiLAM and the reference on eMRS negatives**

|  |  | **Estimate** | **Std. Error** | **z value** | **Pr(>\|z\|)** | **OR** | **2·5 %** | **97·5 %** |
| --- | --- | --- | --- | --- | --- | --- | --- | --- |
| (Intercept) |  | -1·734 | 0·845 | -2·052 | 0·04 | 0·177 | 0·034 | 0·925 |
| ***Age*** |  | 0·002 | 0·01 | 0·252 | 0·801 | 1·002 | 0·983 | 1·022 |
| ***Sex*** | *Male* | 0·583 | 0·231 | 2·524 | 0·012 | 1·792 | 1·139 | 2·818 |
| ***Country*** | *Malawi* | 2·388 | 0·531 | 4·5 | 6·78e-06 | 10·887 | 3·849 | 30·795 |
|  | *Zambia* | 0·956 | 0·558 | 1·714 | 0·087 | 2·602 | 0·872 | 7·766 |
|  | *Uganda* | 0·297 | 0·618 | 0·481 | 0·63 | 1·346 | 0·401 | 4·519 |
|  | *Tanzania* | 0·382 | 0·733 | 0·521 | 0·603 | 1·465 | 0·348 | 6·168 |
|  | *Vietnam* | 0·202 | 0·837 | 0·241 | 0·809 | 1·224 | 0·237 | 6·318 |
|  | *Thailand* | -0·031 | 0·68 | -0·046 | 0·963 | 0·969 | 0·255 | 3·677 |
| ***Lot number*** | *19002* | 1·765 | 0·538 | 3·282 | 0·001 | 5·843 | 2·036 | 16·762 |
|  | *19003* | 0·776 | 0·336 | 2·309 | 0·021 | 2·174 | 1·125 | 4·201 |
|  | *20002* | 3·787 | 1·012 | 3·741 | 1·83e-04 | 44·12 | 6·067 | 320·846 |
|  | *20003* | 4·35 | 0·465 | 9·352 | 8·64e-21 | 77·484 | 31·136 | 192·826 |
|  | *20004* | 2·451 | 0·475 | 5·165 | 2·41e-07 | 11·603 | 4·577 | 29·412 |
| ***Visit day*** | *Day 2* | -0·402 | 0·151 | -2·663 | 0·008 | 0·669 | 0·498 | 0·899 |
| ***CD4*** | *logCD4* | 0·615 | 0·216 | 2·852 | 0·004 | 1·85 | 1·212 | 2·823 |
| ***Urine Color*** | *Clear* | 0·048 | 0·483 | 0·099 | 0·921 | 1·049 | 0·407 | 2·701 |
|  | *Dark yellow* | 0·055 | 0·467 | 0·117 | 0·907 | 1·056 | 0·423 | 2·638 |
|  | *Light yellow* | -0·026 | 0·45 | -0·057 | 0·955 | 0·975 | 0·404 | 2·354 |
|  | *Red* | 14·919 | 882·136 | 0·017 | 0·987 | 3·02e+06 | 0 |  |
| ***Urine Turbidity*** | *Cloudy* | 0·101 | 0·368 | 0·276 | 0·783 | 1·107 | 0·538 | 2·275 |
|  | *Flocculent* | 13·931 | 714·522 | 0·019 | 0·984 | 1·12e+06 | 0 |  |
|  | *Opaque* | 0·417 | 1·025 | 0·407 | 0·684 | 1·518 | 0·203 | 11·327 |
|  | *Slightly cloudy* | 0·21 | 0·226 | 0·928 | 0·353 | 1·234 | 0·792 | 1·923 |
| ***Setting*** | *Outpatient* | -0·868 | 0·311 | -2·791 | 0·005 | 0·42 | 0·228 | 0·772 |

Std., standard; OR, odds ratio.

**Table E18. 111 FujiLAM-positive, eMRS-negative urine samples tested on all six FujiLAM lots and AlereLAM**

| **Lot** | **Positive/All (%)** |
| --- | --- |
| **19001** | 86/111 (77) |
| **19002** | 77/111 (69) |
| **19003** | 78/111 (71) |
| **20002** | 32/111 (29) |
| **20003** | 14/111 (13) |
| **20004** | 14/111 (13) |
| **AlereLAM** | 9/111 (8) |

**Table E19. EclLAM concentration and FujiLAM results from the six lots used in the study using 70 well-characterized urine samples from the FIND specimen bank**

| **Patient TB Category** | **LAM EclLAM (pg/mL)** | **FujiLAM 19001** | **FujiLAM 19002** | **FujiLAM 19003** | **FujiLAM 20002** | **FujiLAM 20003** | **FujiLAM 20004** |
| --- | --- | --- | --- | --- | --- | --- | --- |
| S+C+ | 2031.3 | POS | POS | POS | POS | POS | POS |
| Xp+S-C- | 210.1 | POS | POS | POS | POS | POS | POS |
| S+C+ | 165 | POS | POS | POS | POS | POS | POS |
| S-C+ | 76 | POS | POS | POS | POS | POS | POS |
| S+C+ | 67 | POS | POS | POS | POS | POS | POS |
| S+C+ | 50.5 | POS | POS | POS | POS | POS | POS |
| S+C+ | 46.8 | POS | POS | POS | POS | POS | POS |
| S-C+ | 45 | POS | POS | POS | POS | POS | POS |
| S+C+ | 44.8 | POS | POS | POS | NEG | NEG | POS |
| S-C+ | 41.7 | POS | POS | POS | POS | POS | POS |
| Xp+S-C- | 41.5 | POS | POS | POS | POS | POS | POS |
| S+C+ | 39.8 | POS | POS | POS | POS | POS | POS |
| S+C+ | 37 | POS | POS | POS | POS | POS | POS |
| Xp+S-C- | 34.4 | POS | POS | POS | POS | POS | POS |
| S+C+ | 32.5 | POS | POS | POS | NEG | NEG | NEG |
| S+C+ | 31 | POS | POS | NEG | POS | POS | POS |
| S+C+ | 29.6 | POS | POS | POS | POS | NEG | NEG |
| S+C+ | 29.4 | POS | POS | POS | POS | POS | POS |
| S+C+ | 29 | POS | POS | POS | POS | POS | POS |
| S+C+ | 28.4 | POS | POS | POS | POS | POS | POS |
| S+C+ | 28 | POS | POS | POS | POS | POS | POS |
| S+C+ | 27 | POS | POS | POS | POS | POS | NEG |
| S+C+ | 26.3 | POS | POS | POS | NEG | NEG | NEG |
| S+C+ | 26 | POS | POS | POS | POS | POS | POS |
| S+C+ | 25.1 | POS | POS | POS | POS | NEG | NEG |
| S-C+ | 22.5 | POS | NEG | NEG | NEG | NEG | NEG |
| S+C+ | 22.2 | POS | POS | POS | POS | NEG | NEG |
| S+C+ | 20.5 | POS | POS | POS | NEG | NEG | POS |
| S+C+ | 20 | POS | POS | POS | POS | NEG | POS |
| S+C+ | 19.2 | POS | POS | POS | POS | POS | NEG |
| S-C+ | 19.2 | POS | POS | POS | POS | POS | POS |
| S+C+ | 18.8 | POS | POS | POS | NEG | NEG | NEG |
| S+C+ | 18.6 | POS | POS | POS | POS | NEG | NEG |
| S+C+ | 18 | POS | POS | POS | POS | NEG | POS |
| S+C+ | 17.9 | POS | POS | POS | POS | POS | POS |
| S+C+ | 17.3 | POS | POS | POS | POS | POS | POS |
| S+C+ | 16.3 | POS | POS | POS | POS | NEG | POS |
| S-C+ | 16.1 | POS | POS | POS | POS | POS | POS |
| S+C+ | 15.3 | POS | POS | NEG | NEG | NEG | NEG |
| S+C+ | 14.3 | POS | POS | POS | POS | NEG | POS |
| S+C+ | 13.5 | POS | POS | POS | POS | NEG | POS |
| S-C+ | 12.6 | POS | POS | POS | POS | NEG | POS |
| S+C+ | 12.5 | POS | POS | POS | POS | NEG | NEG |
| S+C+ | 12.4 | POS | POS | POS | NEG | NEG | NEG |
| S+C+ | 11.8 | POS | POS | POS | POS | NEG | POS |
| S+C+ | <LoD | POS | POS | POS | NEG | NEG | NEG |
| S+C+ | <LoD | POS | POS | POS | NEG | NEG | NEG |
| S+C+ | <LoD | POS | POS | POS | NEG | NEG | NEG |
| S+C+ | <LoD | POS | POS | POS | POS | POS | POS |
| S+C+ | <LoD | POS | POS | POS | NEG | NEG | NEG |
| NonTB | <LoD | NEG | POS | POS | NEG | NEG | NEG |
| NonTB | <LoD | POS | POS | POS | NEG | NEG | NEG |
| NonTB | <LoD | POS | POS | NEG | NEG | NEG | NEG |
| NonTB | <LoD | POS | POS | POS | NEG | NEG | NEG |
| NonTB | <LoD | NEG | NEG | NEG | NEG | NEG | NEG |
| NonTB | <LoD | NEG | NEG | NEG | NEG | NEG | NEG |
| NonTB | <LoD | POS | NEG | NEG | NEG | NEG | NEG |
| NonTB | <LoD | POS | POS | POS | NEG | NEG | NEG |
| NonTB | <LoD | POS | POS | POS | NEG | NEG | NEG |
| NonTB | <LoD | POS | POS | POS | NEG | NEG | NEG |
| NonTB | <LoD | POS | POS | POS | NEG | NEG | NEG |
| NonTB | <LoD | POS | POS | POS | NEG | NEG | NEG |
| NonTB | <LoD | POS | POS | POS | NEG | NEG | NEG |
| NonTB | <LoD | POS | POS | POS | NEG | NEG | NEG |
| NonTB | <LoD | POS | POS | POS | POS | NEG | NEG |
| NonTB | <LoD | POS | POS | POS | POS | NEG | NEG |
| NonTB | <LoD | POS | POS | POS | NEG | NEG | NEG |
| NonTB | <LoD | POS | POS | POS | NEG | NEG | NEG |
| NonTB | <LoD | POS | POS | NEG | NEG | NEG | NEG |
| NonTB | <LoD | NEG | POS | NEG | NEG | NEG | NEG |

C, culture; LoD, limit of detection; NEG, negative; NonTB, patients presenting with signs and symptoms suggestive of TB but negative on all available microbiological tests; POS, positive; TB, tuberculosis; S, smear; Xp, Xpert MTB/Rif; S+C+, sputum culture positive and sputum smear microscopy positive; Xp+S-C-, Sputum Xpert MTB/Rif positive, sputum culture and sputum smear microscopy negative.

**Table E20. FujiLAM lots used in previous studies with adult patients**

|  |  |  |  |  | **HIV +** | | **HIV -** | |
| --- | --- | --- | --- | --- | --- | --- | --- | --- |
| **Study** | **Countries** | **Population** | **Overall sample size** | **Lot** | **Sensitivity %** | **Specificity %** | **Sensitivity %** | **Specificity %** |
| **Broger T. et al. Lancet Infect Dis. 2019 (1)** | South Africa | Adult inpatients, HIV+ | 968 | 98002 | 70 | 91 | NA | NA |
| **Bjerrum S. et al. Open Forum Infect Dis. 2019 (2)** | Ghana | Adult in- and outpatients, HIV+ | 532 | 98004 | 74 | 89 | NA | NA |
| **Broger T. et al. PLoS Med. 2020 (3)** | South Africa Ghana  Viet Nam | Adult in- and outpatients, HIV+ | 1595 | 98002 98004 | 70 | 90 | NA | NA |
| **Muyoyeta M. et al. Eur Respir J. 2021 (4)** | Zambia | Adult outpatients, HIV+/- | 151 | 98006 | 77 | 89 | 75 | 95 |
| **Comella-Del-Barrio P. et al. J Clin Med. 2021 (5)** | Nigeria | Adult outpatients, HIV+/- | 204 | 20001 | 70 | 93 | 66 | 99 |
| **Broger T. et al. J Clin Invest. 2020 (6)** | Peru  South Africa | Adult outpatients, HIV- | 372 | 98006 | NA | NA | 53 | 99 |

**Table E21. Evidence to decision tables for FujiLAM lot 19001**

| **Lot 19001** | **Number of results per 1000 patients tested (95% CI)** | | |
| --- | --- | --- | --- |
|  | Prevalence 1% | Prevalence 10% | Prevalence 30% |
| **True positives** | 7 (6 to 8) | 74 (60 to 84) | 222 (180 to 252) |
| **False negatives** | 3 (2 to 4) | 26 (16 to 40) | 78 (48 to 120) |
| **True negatives** | 731 (649 to 766) | 641 (585 to 690) | 499 (455 to 537) |
| **False positives** | 287 (233 to 350) | 259 (210 to 315) | 201 (163 to 245) |

**Table E22. Evidence to decision tables for FujiLAM lot 20003**

| **Lot 20003** | **Number of results per 1000 patients tested (95% CI)** | | |
| --- | --- | --- | --- |
|  | Prevalence 1% | Prevalence 10% | Prevalence 30% |
| **True positives** | 3 (3 to 4) | 34 (26 to 43) | 104 (79 to 132) |
| **False negatives** | 7 (6 to 7) | 66 (57 to 74) | 196 (168 to 221) |
| **True negatives** | 962 (940 to 976) | 867 (847 to 879) | 674 (659 to 684) |
| **False positives** | 37 (23 to 59) | 33 (21 to 53) | 26 (16 to 41) |
